# Prepandemic prevalence estimates of fatty liver disease and fibrosis defined by liver elastography in the United States

**DOI:** 10.1101/2022.04.05.22273458

**Authors:** Aynur Unalp-Arida, Constance E. Ruhl

## Abstract

**Background & Aims:** Fatty liver disease is a growing public health burden with serious consequences. We estimated prepandemic prevalence of fatty liver disease determined by transient elastography assessed hepatic steatosis and fibrosis, and examined associations with lifestyle and other factors in a United States population sample.

**Methods:** Liver stiffness and controlled attenuation parameter (CAP) were assessed on 7,923 non-Hispanic white, non-Hispanic black, non-Hispanic Asian, and Hispanic men and women aged 20 years and over in the National Health and Nutrition Examination Survey (NHANES) 2017-March 2020 prepandemic data.

**Results:** The prevalence of fatty liver disease estimated by CAP >300 dB/m was 28.8% and of fibrosis (liver stiffness >8 kPa) was 10.4%. Only 7.2% of participants with fatty liver disease and 10.9% with fibrosis reported being told by a health care provider that they had liver disease. In addition to known risk factors such as metabolic factors and ALT, persons with fatty liver disease were less likely to meet physical activity guidelines, more likely to be sedentary for 12 or more hours a day, and reported a less healthy diet. Persons with fibrosis were less likely to have a college degree and reported a less healthy diet.

**Conclusion:** In the U.S. population, most persons with fatty liver disease are unaware of their condition. Although physical activity and dietary modifications might reduce the fatty liver disease burden, the COVID pandemic has been less favorable for lifestyle changes. There is an urgent need for fatty liver disease management in high-risk individuals using transient elastography or other noninvasive methods to intervene in disease progression.

## INTRODUCTION

Fatty liver disease is responsible for considerable morbidity, mortality, and economic cost and is a growing public health burden.^1^ Chronic liver disease and cirrhosis lead to increased morbidity and ranked as the 11^th^ leading cause of death in the United States in 2019.^2^ Fatty liver disease is a leading cause of chronic liver disease in the U.S. and similar to nonalcoholic fatty liver disease (NAFLD), it can progress to severe fibrosis with increased risk of liver-related complications and death.^3,4^ The burden of fatty liver disease is expected to grow over the next decade due to lingering pandemic effects, the worsening epidemic of obesity and diabetes in the absence of widespread public health measures, and the lack of optimal care of the growing number of patients with NAFLD.^5,6^

NAFLD is considered part of a broader multi-system disease that also includes obesity, diabetes, hyperlipidemia, and hypertension. Consequently, redefining NAFLD as metabolic dysfunction-associated fatty liver disease (MAFLD) based on modified criteria was recently proposed.^7^ However, there are few population data on the epidemiologic implications of this redefinition.^8-10^

Lifestyle modifications are the first line of treatment for fatty liver disease based on its weight-related etiology and the efficacy of weight loss in reversing disease progression.^11^ Unhealthy diets and physical inactivity have been linked to higher fatty liver disease risk and hence, dietary modifications and increased physical activity may be effective in its management. Increased physical activity is recommended along with a low calorie diet to achieve weight loss among persons with fatty liver disease despite limited information on the relationship of physical activity and diet with fatty liver disease in the general U.S. population.^12-14^ Recently, the COVID-19 pandemic has augmented health disparities and imposes further challenges in implementation of these lifestyle improvements. A better understanding of social determinants of health, such as access to healthy food or places to exercise, may help to reduce health disparities and achieve health equity which may decrease the fatty liver disease burden among racial and ethnic minority and other underserved populations.^15^

Transient elastography by FibroScan^®^ is a practical noninvasive tool for tracking liver health in the general population and has been widely used for chronic liver disease surveillance over the past decade.^16-17^ However, population-based data on prevalence of transient elastography assessed fatty liver disease and fibrosis in the United States and associated factors are limited.^18-19^ Transient elastography was used to measure hepatic steatosis and fibrosis beginning in NHANES 2017-2018 which provided the first U.S. representative transient elastography liver stiffness and CAP observations.^20^ Using those data, we reported transient elastography-derived hepatic steatosis and fibrosis distributions in U.S. adults and their associations with body composition.^21^ That analysis was limited by the smaller sample size with available transient elastography data using a single 2-year survey cycle. Additional liver elastography data recently became available with the release of the NHANES 2017-March 2020 prepandemic data enabling increased precision of prevalence estimates and ability to detect differences among population groups to further explain liver disease heterogeneity. In the current report, we estimated prepandemic prevalence of fatty liver disease determined by transient elastography assessed hepatic steatosis and fibrosis, and of NAFLD and MAFLD in a United States population sample. We also examined fatty liver disease associations, including those with lifestyle factors not available at the time of our previous analysis, to investigate the potential impact of adverse social factors.

## MATERIALS AND METHODS

### Source population

The NHANES is conducted in the United States by the National Center for Health Statistics (NCHS) of the Centers for Disease Control and Prevention (CDC).^22^ The survey consists of interview, examination, and laboratory data collected from a complex multistage, stratified, clustered probability sample representative of the civilian, noninstitutionalized population with oversampling of non-Hispanic blacks, Hispanics, Asians, low income persons, and older adults. The CDC ethics review board approved the survey, and all participants provided written informed consent. The current analysis utilized data collected from 2017 through March 2020 when the CDC NCHS stopped the survey due to the emerging COVID-19 pandemic, during which transient elastography was used to measure hepatic steatosis and fibrosis.

### Transient elastography

Vibration controlled transient elastography using FibroScan^®^ 502 V2 Touch (Echosens™ North America, Waltham, MA) was performed on eligible NHANES 2017-March 2020 participants.^20^ Multiple measurements up to ≥30 were made on each participant using a medium (M; 72% of participants) or large (XL) probe. Hepatic fibrosis was measured using transient elastography-derived liver stiffness in kilopascals (kPa) with median, interquartile range (IQR), and IQR/median calculated for each participant. Simultaneously, hepatic steatosis was measured using CAP in decibels per meter (dB/m) with median and IQR calculated for each participant.

### Other variables

Age (years), sex, race-ethnicity (non-Hispanic white, non-Hispanic black, non-Hispanic Asian, Hispanic, or other), education (less than high school graduate, high school graduate or GED or equivalent, some college or associate degree, college graduate or above), income, diagnosed diabetes, blood pressure medication use, liver disease history, cigarette smoking (never, former, current), alcohol use, health insurance coverage (private, public only, or uninsured), physical activity, and healthiness of diet (excellent/very good, good, or fair/poor) were ascertained by interview. Income was measured by the poverty income ratio (ratio of family income to poverty threshold) and categorized as quartiles. Current alcohol use was categorized as none, moderate (>0-<3 drinks/day for men or >0-<2 drink/day for women), or heavy (≥3 drinks/day for men or ≥2 drink/day for women). Whether or not the participant met WHO physical activity guidelines and sedentary activity (hours per day) were determined from responses to the Global Physical Activity Questionnaire.^23^ A person was considered to have met the guidelines if they did at least 150-300 minutes of moderate-intensity aerobic physical activity, or at least 75-150 minutes of vigorous-intensity aerobic physical activity, or an equivalent combination of moderate-and vigorous-intensity activity per week.

Body measurements including standing height (cm), weight (kg), waist circumference (cm), and hip circumference (cm) were ascertained during the mobile examination center visit, and body mass index (BMI; weight [kg]/height [m^2^]) and waist-to-hip circumference ratio were calculated. Systolic and diastolic blood pressure (mmHg) were measured. Serum was tested for hemoglobin A_1C_ (%), total and high-density lipoprotein (HDL) cholesterol (mg/dL), alanine aminotransferase (ALT, IU/L), aspartate aminotransferase (AST, IU/L), gamma-glutamyltransferase (GGT, IU/L), and high-sensitivity C-reactive protein (mg/L). Among participants examined in the morning after an overnight fast of 8 to less than 24 hours, serum was also tested for triglycerides (mg/dL), glucose (mg/dL), and insulin (pmol/L). Pre-diabetes was defined as hemoglobin A_1C_ 5.7%-<6.5% or fasting glucose (mg/dL) 100-<126 and diabetes as a health care provider diagnosis, hemoglobin A_1C_ ≥6.5%, or fasting glucose ≥126 mg/dL. Elevated blood pressure was defined as systolic blood pressure ≥130 mmHg, diastolic blood pressure ≥80 mmHg or blood pressure medication use.

Fatty liver disease was defined as CAP >300 dB/m and fibrosis as liver stiffness >8 kPa based on the optimized sensitivity and specificity reported in earlier studies.^24-26^ NAFLD was identified by CAP >300 dB/m among men with alcohol use of <3 drinks per day or women with <2 drinks per day.^27^ MAFLD was defined as CAP >300 dB/m among participants with overweight or obese (BMI ≥23 kg/m^2^ among non-Hispanic Asians or ≥25 kg/m^2^ among non-Asians), diabetes, or at least two metabolic risk abnormalities.^7^ Metabolic risk abnormalities consisted of waist circumference ≥90 cm for men or ≥80 cm for women among non-Hispanic Asians or ≥102 cm for men or ≥88 cm for women among non-Asians, blood pressure ≥130/85 mmHg or blood pressure medication use, fasting triglycerides ≥150 mg/dL, HDL cholesterol <40 mg/dL for men or <50 mg/dL for women, prediabetes, homeostasis model assessment of insulin resistance score ≥2.5, or high-sensitivity C-reactive protein >2 mg/L.

### Analysis sample

Of 9,232 NHANES 2017-March 2020 participants 20 years and older who were interviewed at home, 8,544 attended a mobile examination center visit. Persons ineligible for liver elastography (pregnant, undetermined pregnancy status, insulin pump or other implantable electronic device) were excluded (n=333). Participants who did not undergo liver elastography (refused, insufficient time, physical or technical limitations) or had no complete stiffness measures were also excluded (n=288). The resulting liver elastography analysis sample consisted of 7,923 persons (**Supplemental Figure 1**). Of these, 7,396 (93.6%) had complete liver elastography and 527 were considered by the NCHS to have a partial examination because of fasting < 3 hours (n=248), 1 to <10 valid measures obtained (n=99), or a stiffness IQR/median ≥ 30% (n=180) **(Supplemental Table 1)**. Higher BMI, stiffness, and CAP of participants in the latter two groups suggested they had potentially more severe liver injury; consequently, as in our previous NHANES 2017-2018 analysis we included them in our current analysis to reflect the full range of U.S. representative population liver disease.^21^ Analyses including fasting laboratory measures were conducted among morning examined fasted participants (n=3,576).

### Statistical analysis

We evaluated characteristics of participants with complete and partial liver elastography by comparing continuous factor means (standard deviations (SD)) using analysis of variance and categorical factor percentages using chi-square (χ^2^) tests or linear regression. Unadjusted relationships of fatty liver disease and fibrosis with demographic, clinical, lifestyle, and social characteristics were examined by comparing continuous factor means (SD) using t-tests and categorical factor percentages using chi-square (χ^2^) tests. Age-adjusted relationships were explored using logistic regression to calculate odds ratios (OR) and 95% confidence intervals (CI). Multiple logistic regression analysis was used to determine relationships of fatty liver disease and fibrosis with participant characteristics after adjustment for all other factors in the model. Age, sex, and race-ethnicity were retained in models. A biological plausibility approach was followed to test anthropometric measures (BMI, BMI and waist-to-hip ratio, and waist circumference), metabolic factors (diabetes, total and HDL cholesterol, elevated blood pressure, C-reactive protein, and CAP (for liver stiffness models)), liver enzymes (ALT, AST, and GGT), lifestyle factors (alcohol use, smoking, physical activity, and diet), and social factors (education, income, and health insurance coverage) for inclusion in models.^28^ For factors that were highly correlated, the measure that explained the greatest variance (R^2^) was selected. Factors were retained in final models if p<0.10. Factors that were not normally distributed were expressed as deciles (10^th^ percentiles) for regression analyses using cut points shown in **Supplemental Table 2**. P-values were two-sided, and P<0.05 indicated statistical significance. Sample weights accounted for unequal selection probabilities and nonresponse. Variance calculations accounted for survey design effects using Taylor series linearization.^29^ SAS 9.4 (SAS Institute, Cary, NC) and SUDAAN 11 (RTI, Research Triangle Park, NC) were used.

## RESULTS

Among 7,923 adult participants with liver elastography measures, mean (SD) CAP was 264.6 (63.1) dB/m and mean liver stiffness was 5.9 (4.9) kPa (**Table 1**). The overall prevalence of fatty liver disease (CAP >300 dB/m) was 28.8% (95% C.I., 27.1%-30.6%). The prevalence of fibrosis risk (liver stiffness >8 kPa) was 10.4% (95% C.I., 8.8%-12.2%) in the overall population and as high as 22.8% (95% C.I., 19.2%-26.8%) among persons with fatty liver disease. The NAFLD definition (CAP >300 dB/m without heavy alcohol use) was met by 26.4% (95% C.I., 24.7%-28.2%) of participants, while 1.3% had elevated CAP with heavy alcohol use and 1.1% had elevated CAP and missing alcohol use data so NAFLD status could not be determined. The MAFLD definition (CAP >300 dB/m and fulfilled metabolic criteria) was met by 28.3% (95% C.I., 26.6%-30.1%) of participants, while 0.4% had elevated CAP, but did not fulfill metabolic criteria and 0.1% had elevated CAP, but MAFLD status could not be determined due to incomplete metabolic data. Prevalence of fibrosis did not differ between persons with NAFLD (22.9%; 95% C.I., 19.2%-27.0%) and MAFLD (22.9%; 95% C.I., 19.3%-27.1%).

**Table 1.**
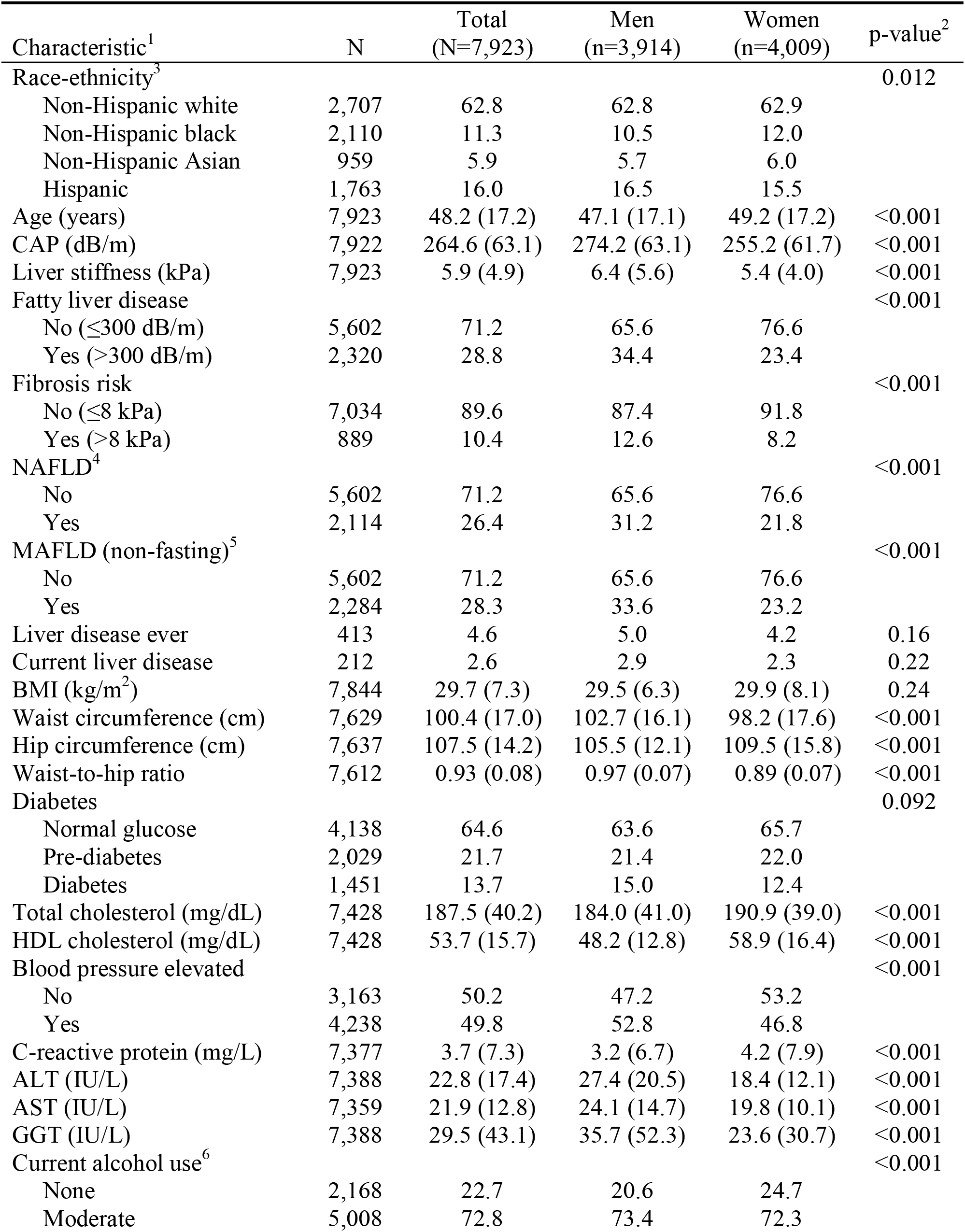
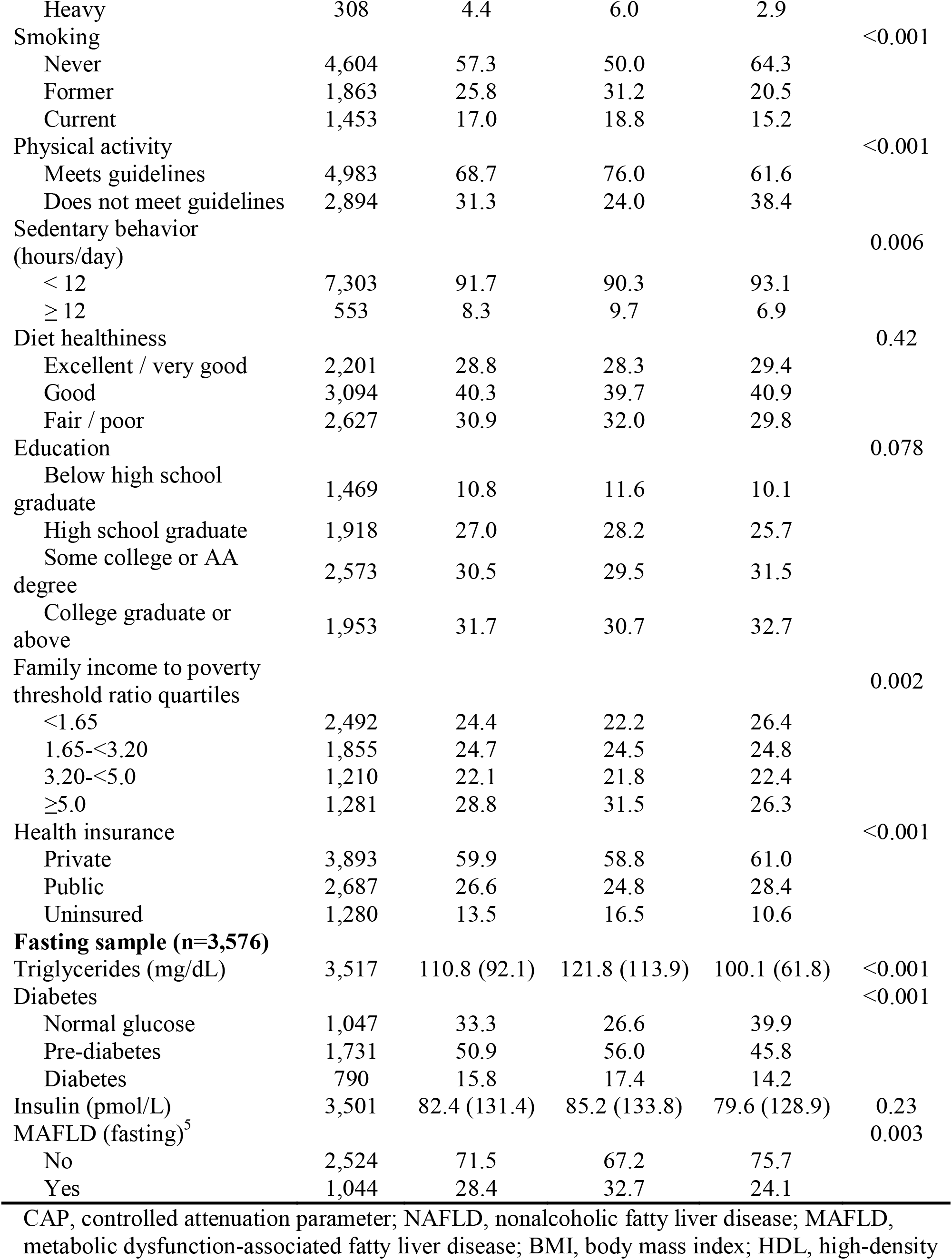
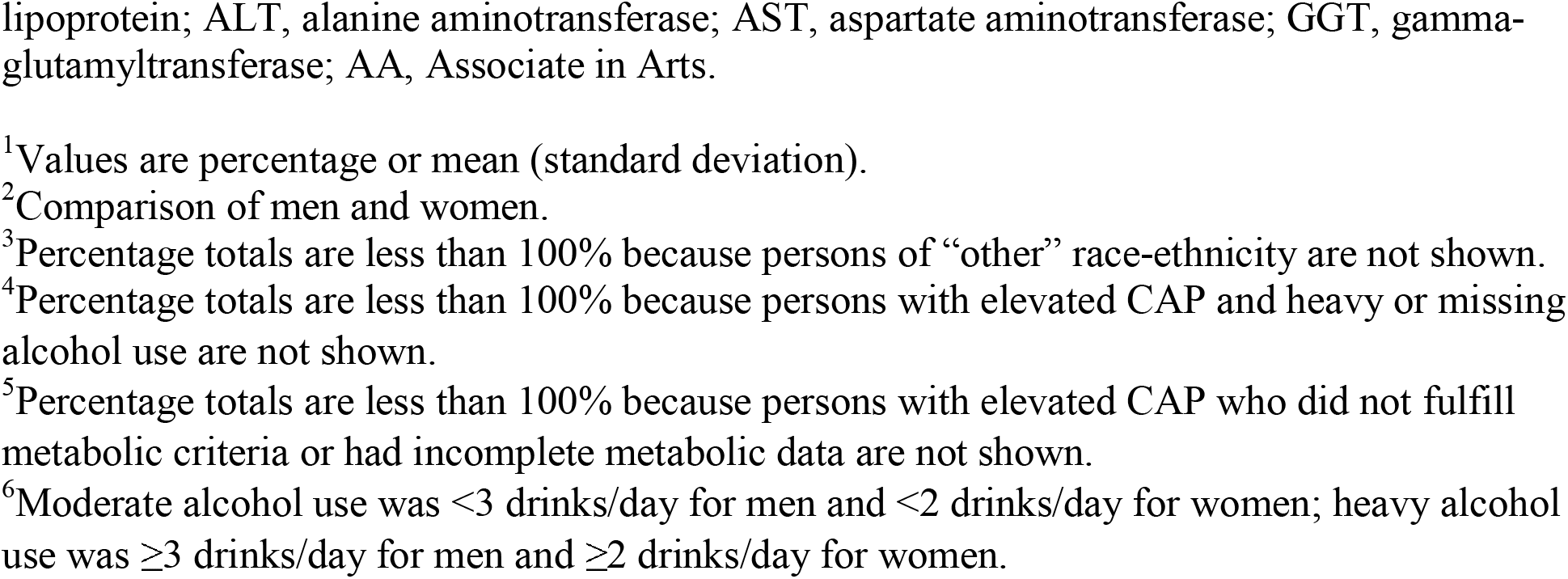
Characteristics of participants 20+ years in the National Health and Nutrition Examination Survey, United States, 2017-March 2020 prepandemic data

Health care provider diagnosed liver disease ever was reported by 4.6% of all participants and a current liver disease diagnosis by 2.6%. The mean (SD) duration of liver disease was 11.0 (12.3) years. The prevalence was 2.3% for fatty liver (in contrast to transient elastography assessed 28.8%), 0.1% for liver fibrosis (in contrast to transient elastography assessed 10.4%), 0.3% for liver cirrhosis, 0.8% for viral hepatitis, 0.2% for autoimmune hepatitis, and 1.2% for other liver disease. Men had a higher prevalence of transient elastography identified fatty liver disease, fibrosis risk, NAFLD, and MAFLD compared with women **(Table 1)**. Sex differences in other participant characteristics were similar to those reported previously among the NHANES 2017-2018 sample.^21^ In addition, while women less often met physical activity guidelines, they were also less likely to spend 12 or more hours per day in sedentary behavior. Among the fasting sample, men had higher triglyceride and glucose levels.

### Fatty liver disease

Participants with fatty liver disease were more likely to be Hispanic and less likely to be non-Hispanic black or non-Hispanic Asian **(Table 2)**. This was consistent among both men and women (**Figure 1**). Among men, fatty liver disease prevalence was higher among non-Hispanic Asians compared with non-Hispanic blacks; whereas, among women non-Hispanics blacks and non-Hispanic Asians did not differ significantly. Among persons with fatty liver disease, only 7.2% reported ever having been told by a health care provider that they had liver disease and only 4.3% reported still having liver disease. Persons with fatty liver disease were older and more likely to have metabolic risk factors, higher liver enzymes, and to be former smokers. They were less likely to meet physical activity guidelines, more likely to be sedentary for 12 or more hours a day, and self-reported a less healthy diet **(Table 2, Figure 2A)**. Fatty liver disease prevalence was highest among persons who did not meet physical activity guidelines and were sedentary for 12 or more hours a day **(Figure 2B)**. The higher fatty liver disease prevalence among persons with less favorable lifestyle habits was seen across sex and race-ethnicity groups, although differences did not reach statistical significance among all groups (**Supplemental Figures 2-6)**. Participants with fatty liver disease were less likely to have a college degree, but did not differ with regard to income **(Table 2, Supplemental Figure 7)**. The lower fatty liver disease prevalence among persons with the highest education level was seen across sex and race-ethnicity groups, although differences did not reach statistical significance among all groups (**Supplemental Figures 8-11)**. ALT, AST, and GGT were elevated among only 22.7%, 10.7%, and 22.5% of persons with fatty liver disease, respectively **(Table 2)**.

**Table 2.** Unadjusted associations with fatty liver disease and fibrosis risk among participants 20+ years in the National Health and Nutrition Examination Survey, United States, 2017-March 2020 prepandemic data (N=7,923)

| Characteristic<sup>1</sup> | No fatty liver disease (≤300 dB/m) | Fatty liver disease (>300 dB/m) | No fibrosis risk (≤8 kPa) | Fibrosis risk (>8 kPa) |
| --- | --- | --- | --- | --- |
| Women | 54.7 | 41.3<sup>a</sup> | 52.1 | 40.2<sup>a</sup> |
| Race-ethnicity<sup>2</sup> |  |  |  |  |
| Non-Hispanic white | 62.6 | 63.5 | 62.5 | 65.8 |
| Non-Hispanic black | 12.5 | 8.2<sup>a</sup> | 11.4 | 10.4 |
| Non-Hispanic Asian | 6.2 | 5.0<sup>b</sup> | 6.1 | 4.0<sup>b</sup> |
| Hispanic | 14.8 | 19.0<sup>b</sup> | 16.1 | 15.3 |
| Age (years) | 47.0 (17.5) | 51.0 (16.0)<sup>a</sup> | 47.7 (17.2) | 51.9 (16.1)<sup>a</sup> |
| CAP (dB/m) | 233.0 (42.1) | 342.4 (30.4)<sup>a</sup> | 258.7 (60.3) | 315.4 (63.8)<sup>a</sup> |
| Liver stiffness (kPa) | 5.2 (3.5) | 7.6 (7.0)<sup>a</sup> | 4.9 (1.2) | 14.6 (11.5)<sup>a</sup> |
| Fatty liver disease (>300 dB/m) | -- | -- | 24.8 | 63.2<sup>a</sup> |
| Fibrosis risk (>8 kPa) | 5.4 | 22.8<sup>a</sup> |  |  |
| Liver disease ever | 3.6 | 7.2<sup>a</sup> | 3.9 | 10.9<sup>b</sup> |
| Current liver disease | 1.9 | 4.3<sup>a</sup> | 2.0 | 8.1<sup>b</sup> |
| BMI (kg/m<sup>2</sup>) | 27.5 (5.9) | 35.1 (7.5)<sup>a</sup> | 28.9 (6.4) | 36.5 (9.9)<sup>a</sup> |
| Waist circumference (cm) | 94.7 (14.2) | 114.4 (15.3)<sup>a</sup> | 98.6 (15.6) | 116.8 (20.2)<sup>a</sup> |
| Hip circumference (cm) | 103.8 (11.9) | 116.6 (15.3)<sup>a</sup> | 106.2 (12.9) | 119.3 (19.2)<sup>a</sup> |
| Waist-to-hip ratio | 91.1 (7.6) | 98.4 (7.2)<sup>a</sup> | 92.7 (8.0) | 98.2 (7.9)<sup>a</sup> |
| Pre-diabetes | 18.8 | 28.7<sup>a</sup> | 21.2 | 25.6 |
| Diabetes | 8.0 | 27.8<sup>a</sup> | 11.2 | 35.9<sup>a</sup> |
| Total cholesterol (mg/dL) | 186.5 (39.7) | 190.0 (41.0)<sup>a</sup> | 188.2 (40.0) | 181.6 (41.2)<sup>a</sup> |
| HDL cholesterol (mg/dL) | 56.5 (15.7) | 46.7 (13.2)<sup>a</sup> | 54.2 (15.7) | 48.6 (14.9)<sup>a</sup> |
| Blood pressure elevated | 42.6 | 67.2<sup>a</sup> | 46.8 | 76.0<sup>a</sup> |
| C-reactive protein (mg/L) | 3.1 (7.1) | 5.2 (7.6)<sup>a</sup> | 3.5 (7.1) | 5.6 (9.0)<sup>a</sup> |
| ALT (IU/L) | 20.0 (15.0) | 29.7 (20.6)<sup>a</sup> | 21.7 (15.2) | 33.1 (28.3)<sup>a</sup> |
| ALT elevated<sup>3</sup> | 6.6 | 22.7<sup>a</sup> | 9.3 | 28.4<sup>a</sup> |
| AST (IU/L) | 20.9 (12.1) | 24.2 (13.9)<sup>a</sup> | 21.0 (9.9) | 29.5 (25.7)<sup>a</sup> |
| AST elevated<sup>4</sup> | 4.6 | 10.7<sup>a</sup> | 4.8 | 19.7<sup>a</sup> |
| GGT (IU/L) | 25.7 (42.2) | 39.0 (43.8)<sup>a</sup> | 26.5 (29.2) | 55.8 (99.4)<sup>a</sup> |
| GGT elevated<sup>5</sup> | 9.9 | 22.5<sup>a</sup> | 11.2 | 34.2<sup>a</sup> |
| Moderate alcohol use<sup>6</sup> | 74.0 | 70.2 | 73.5 | 67.4<sup>b</sup> |
| Heavy alcohol use<sup>6</sup> | 4.4 | 4.6 | 4.4 | 4.9 |
| Former smoker | 23.4 | 31.6<sup>a</sup> | 25.1 | 31.8<sup>b</sup> |
| Current smoker | 17.4 | 15.8 | 17.0 | 16.4 |
| Physical inactivity<sup>7</sup> | 28.3 | 38.6<sup>a</sup> | 30.2 | 40.8<sup>a</sup> |
| Sedentary behavior, ≥12 hours/day | 7.0 | 11.4<sup>b</sup> | 7.9 | 11.9<sup>b</sup> |
| Diet healthiness |  |  |  |  |
| Excellent / very good | 32.5 | 19.7<sup>a</sup> | 30.0 | 18.5<sup>a</sup> |
| Good | 40.9 | 38.9 | 40.5 | 38.8 |
| Fair / poor | 26.6 | 41.3<sup>a</sup> | 29.5 | 42.6<sup>a</sup> |
| Education |  |  |  |  |
| < high school graduate | 10.7 | 11.2 | 10.8 | 11.4 |
| High school graduate | 25.7 | 30.0<sup>b</sup> | 25.9 | 35.8<sup>b</sup> |
| Some college / AA degree | 29.2 | 33.6<sup>b</sup> | 30.3 | 31.9 |
| College graduate or above | 34.3 | 25.3<sup>a</sup> | 33.0 | 20.9<sup>a</sup> |
| Family income to poverty threshold ratio quartiles |  |  |  |  |
| <1.65 | 23.9 | 25.6 | 24.3 | 25.2 |
| 1.65-<3.20 | 24.0 | 26.4 | 23.7 | 32.7<sup>b</sup> |
| 3.20-<5.0 | 22.0 | 22.5 | 22.5 | 19.2 |
| ≥5.0 | 30.2 | 25.5 | 29.5 | 22.9<sup>b</sup> |
| Health insurance |  |  |  |  |
| Private | 60.2 | 59.2 | 60.0 | 58.9 |
| Public | 25.7 | 28.9 | 26.3 | 29.1 |
| Uninsured | 14.2 | 11.9<sup>b</sup> | 13.6 | 12.1 |
| **Fasting sample (n=3,576)** |  |  |  |  |
| Triglycerides (mg/dL) | 94.9 (78.4) | 150.5 (110.0)<sup>a</sup> | 108.9 (89.6) | 128.7 (111.5)<sup>a</sup> |
| Pre-diabetes | 50.9 | 50.8 | 51.7 | 43.2 |
| Diabetes | 8.9 | 33.1<sup>a</sup> | 13.0 | 42.4<sup>a</sup> |
| Insulin (pmol/L) | 61.3 (95.8) | 134.8 (183.2)<sup>a</sup> | 73.4 (79.3) | 166.1 (334.5)<sup>b</sup> |

CAP, controlled attenuation parameter; BMI, body mass index; HDL, high-density lipoprotein; ALT, alanine aminotransferase; AST, aspartate aminotransferase; GGT, gamma-glutamyltransferase; AA, Associate in Arts.
<sup>1</sup>Values are percentage or mean (standard deviation).
<sup>2</sup>Percentage total is less than 100% because persons of “other” race-ethnicity are not shown.
<sup>3</sup>ALT >40 IU/L in men or >31 IU/L in women.
<sup>4</sup>AST >37 IU/L in men or >31 IU/L in women.
<sup>5</sup>GGT >51 IU/L in men or >33 IU/L in women.
<sup>6</sup>Moderate alcohol use was <3 drinks/day for men and <2 drinks/day for women; heavy alcohol use was ≥3 drinks/day for men and ≥2 drinks/day for women.
<sup>7</sup>Does not meet physical activity guidelines.
Compared to fatty liver disease or fibrosis risk: <sup>a</sup>p<0.001, <sup>b</sup>p<0.05.

**Figure 1.**
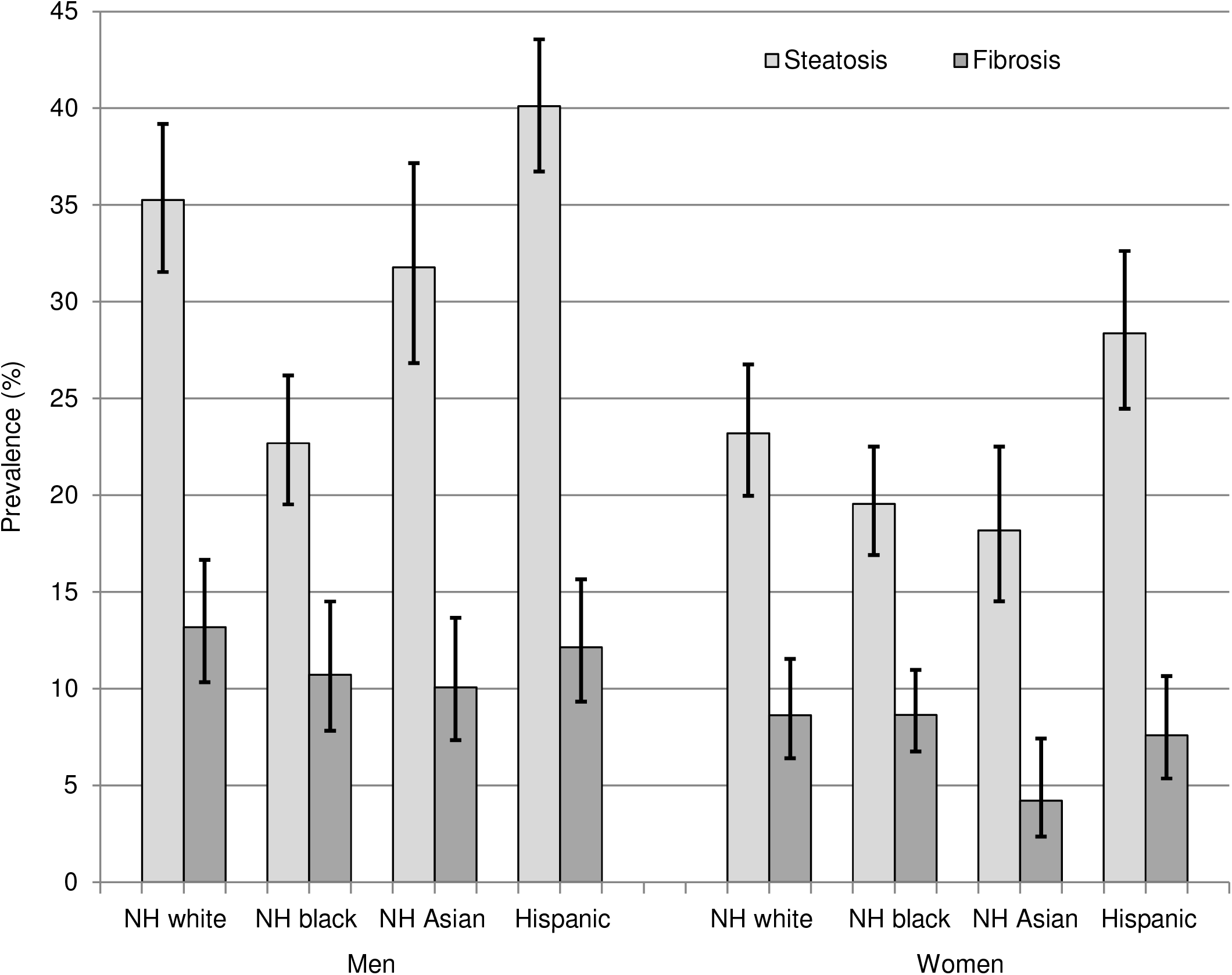
Prevalence of fatty liver disease and fibrosis by sex and race-ethnicity among participants 20+ years in the National Health and Nutrition Examination Survey, United States, 2017-March 2020 prepandemic data (N=7,923) NH, non-Hispanic.

**Figure 2.**
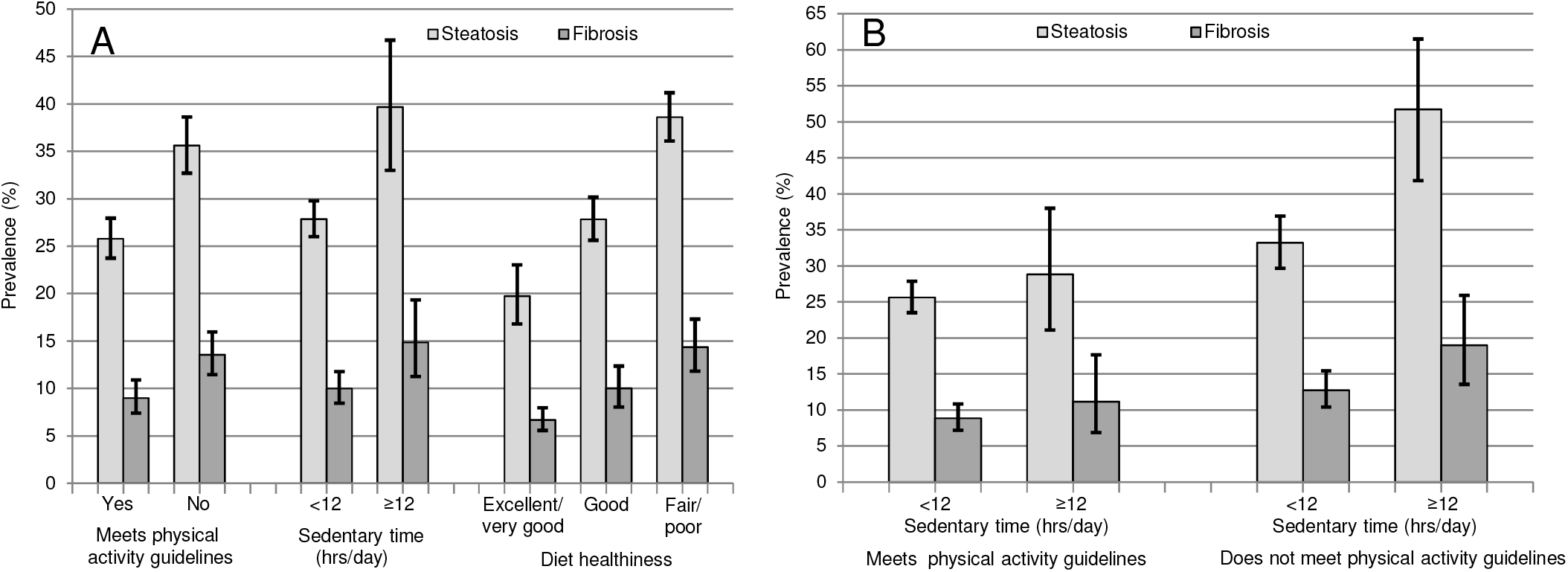
Prevalence of fatty liver disease and fibrosis among participants 20+ years in the National Health and Nutrition Examination Survey, United States, 2017-March 2020 prepandemic data (N=7,923) (**A)**. By lifestyle factors. (**B)**. By physical activity and sedentary time.

With adjustment for age, non-Hispanic Asian race-ethnicity was no longer inversely associated with fatty liver disease (**Table 3**). An association appeared with the lowest income quartile, while other relationships were similar. In multivariate-adjusted analysis, fatty liver disease was positively associated with non-Hispanic Asian race-ethnicity (**Table 4**). Associations remained with older age, prediabetes and diabetes, higher BMI, waist-to-hip ratio, total cholesterol, blood pressure, C-reactive protein, and ALT and inversely with non-Hispanic black race-ethnicity and HDL cholesterol. There was a trend toward an association with less physical activity, more sedentary behavior, and a less healthy diet (0.05<p<0.1). Education and income were no longer associated. Among persons who fasted, factors most strongly associated with fatty liver disease in multivariate-adjusted analysis were non-Hispanic Asian race-ethnicity, diabetes, higher BMI, waist-to-hip ratio, triglycerides, insulin, total cholesterol, ALT, and sedentary time, and an inverse association with non-Hispanic black race-ethnicity (**Table 5**). Because of the smaller sample size compared with **Table 4** and additional adjustment for triglycerides and insulin, some factors from the full sample model no longer remained associated (i.e. pre-diabetes, HDL cholesterol, elevated blood pressure, C-reactive protein, physical activity, and diet). In contrast, sedentary behavior became more strongly associated with fatty liver.

**Table 3.**
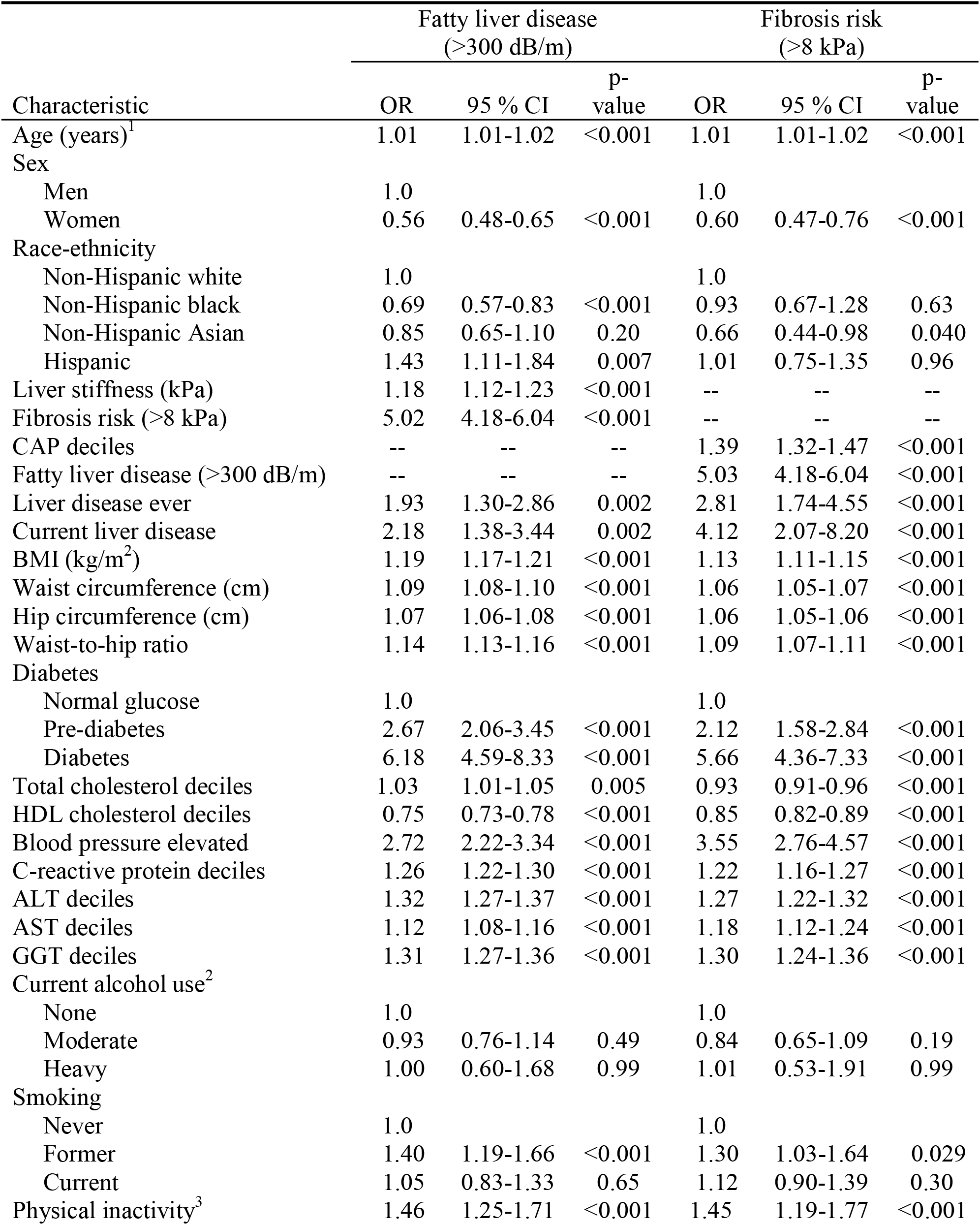
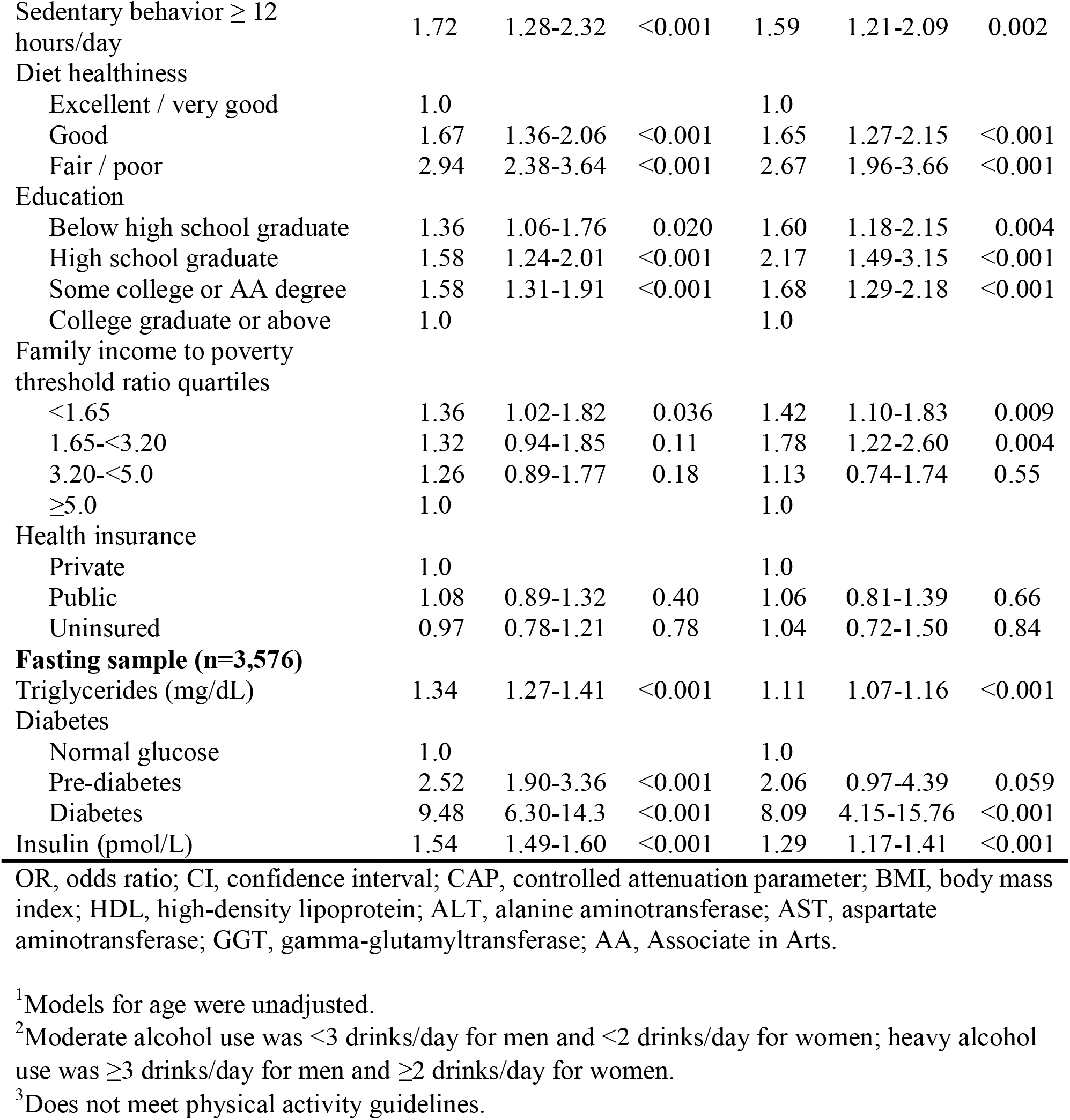
Age-adjusted logistic regression associations of fatty liver disease and fibrosis risk among participants 20+ years in the National Health and Nutrition Examination Survey, United States, 2017-March 2020 prepandemic data (N=7,923)

| Characteristic | Fatty liver disease<br>(>300 dB/m) |  |  | Fibrosis risk<br>(>8 kPa) |  |  |
| --- | --- | --- | --- | --- | --- | --- |
|  | OR | 95 % CI | p-value | OR | 95 % CI | p-value |
| Age (years)<sup>1</sup> | 1.01 | 1.01-1.02 | <0.001 | 1.01 | 1.01-1.02 | <0.001 |
| Sex |  |  |  |  |  |  |
| Men | 1.0 |  |  | 1.0 |  |  |
| Women | 0.56 | 0.48-0.65 | <0.001 | 0.60 | 0.47-0.76 | <0.001 |
| Race-ethnicity |  |  |  |  |  |  |
| Non-Hispanic white | 1.0 |  |  | 1.0 |  |  |
| Non-Hispanic black | 0.69 | 0.57-0.83 | <0.001 | 0.93 | 0.67-1.28 | 0.63 |
| Non-Hispanic Asian | 0.85 | 0.65-1.10 | 0.20 | 0.66 | 0.44-0.98 | 0.040 |
| Hispanic | 1.43 | 1.11-1.84 | 0.007 | 1.01 | 0.75-1.35 | 0.96 |
| Liver stiffness (kPa) | 1.18 | 1.12-1.23 | <0.001 | -- | -- | -- |
| Fibrosis risk (>8 kPa) | 5.02 | 4.18-6.04 | <0.001 | -- | -- | -- |
| CAP deciles | -- | -- | -- | 1.39 | 1.32-1.47 | <0.001 |
| Fatty liver disease (>300 dB/m) | -- | -- | -- | 5.03 | 4.18-6.04 | <0.001 |
| Liver disease ever | 1.93 | 1.30-2.86 | 0.002 | 2.81 | 1.74-4.55 | <0.001 |
| Current liver disease | 2.18 | 1.38-3.44 | 0.002 | 4.12 | 2.07-8.20 | <0.001 |
| BMI (kg/m<sup>2</sup>) | 1.19 | 1.17-1.21 | <0.001 | 1.13 | 1.11-1.15 | <0.001 |
| Waist circumference (cm) | 1.09 | 1.08-1.10 | <0.001 | 1.06 | 1.05-1.07 | <0.001 |
| Hip circumference (cm) | 1.07 | 1.06-1.08 | <0.001 | 1.06 | 1.05-1.06 | <0.001 |
| Waist-to-hip ratio | 1.14 | 1.13-1.16 | <0.001 | 1.09 | 1.07-1.11 | <0.001 |
| Diabetes |  |  |  |  |  |  |
| Normal glucose | 1.0 |  |  | 1.0 |  |  |
| Pre-diabetes | 2.67 | 2.06-3.45 | <0.001 | 2.12 | 1.58-2.84 | <0.001 |
| Diabetes | 6.18 | 4.59-8.33 | <0.001 | 5.66 | 4.36-7.33 | <0.001 |
| Total cholesterol deciles | 1.03 | 1.01-1.05 | 0.005 | 0.93 | 0.91-0.96 | <0.001 |
| HDL cholesterol deciles | 0.75 | 0.73-0.78 | <0.001 | 0.85 | 0.82-0.89 | <0.001 |
| Blood pressure elevated | 2.72 | 2.22-3.34 | <0.001 | 3.55 | 2.76-4.57 | <0.001 |
| C-reactive protein deciles | 1.26 | 1.22-1.30 | <0.001 | 1.22 | 1.16-1.27 | <0.001 |
| ALT deciles | 1.32 | 1.27-1.37 | <0.001 | 1.27 | 1.22-1.32 | <0.001 |
| AST deciles | 1.12 | 1.08-1.16 | <0.001 | 1.18 | 1.12-1.24 | <0.001 |
| GGT deciles | 1.31 | 1.27-1.36 | <0.001 | 1.30 | 1.24-1.36 | <0.001 |
| Current alcohol use<sup>2</sup> |  |  |  |  |  |  |
| None | 1.0 |  |  | 1.0 |  |  |
| Moderate | 0.93 | 0.76-1.14 | 0.49 | 0.84 | 0.65-1.09 | 0.19 |
| Heavy | 1.00 | 0.60-1.68 | 0.99 | 1.01 | 0.53-1.91 | 0.99 |
| Smoking |  |  |  |  |  |  |
| Never | 1.0 |  |  | 1.0 |  |  |
| Former | 1.40 | 1.19-1.66 | <0.001 | 1.30 | 1.03-1.64 | 0.029 |
| Current | 1.05 | 0.83-1.33 | 0.65 | 1.12 | 0.90-1.39 | 0.30 |
| Physical inactivity<sup>3</sup> | 1.46 | 1.25-1.71 | <0.001 | 1.45 | 1.19-1.77 | <0.001 |
| Sedentary behavior $\geq 12$ hours/day | 1.72 | 1.28-2.32 | <0.001 | 1.59 | 1.21-2.09 | 0.002 |
| Diet healthiness |  |  |  |  |  |  |
| Excellent / very good | 1.0 |  |  | 1.0 |  |  |
| Good | 1.67 | 1.36-2.06 | <0.001 | 1.65 | 1.27-2.15 | <0.001 |
| Fair / poor | 2.94 | 2.38-3.64 | <0.001 | 2.67 | 1.96-3.66 | <0.001 |
| Education |  |  |  |  |  |  |
| Below high school graduate | 1.36 | 1.06-1.76 | 0.020 | 1.60 | 1.18-2.15 | 0.004 |
| High school graduate | 1.58 | 1.24-2.01 | <0.001 | 2.17 | 1.49-3.15 | <0.001 |
| Some college or AA degree | 1.58 | 1.31-1.91 | <0.001 | 1.68 | 1.29-2.18 | <0.001 |
| College graduate or above | 1.0 |  |  | 1.0 |  |  |
| Family income to poverty threshold ratio quartiles |  |  |  |  |  |  |
| <1.65 | 1.36 | 1.02-1.82 | 0.036 | 1.42 | 1.10-1.83 | 0.009 |
| 1.65-<3.20 | 1.32 | 0.94-1.85 | 0.11 | 1.78 | 1.22-2.60 | 0.004 |
| 3.20-<5.0 | 1.26 | 0.89-1.77 | 0.18 | 1.13 | 0.74-1.74 | 0.55 |
| $\geq 5.0$ | 1.0 | | | 1.0 | | |
| Health insurance |  |  |  |  |  |  |
| Private | 1.0 |  |  | 1.0 |  |  |
| Public | 1.08 | 0.89-1.32 | 0.40 | 1.06 | 0.81-1.39 | 0.66 |
| Uninsured | 0.97 | 0.78-1.21 | 0.78 | 1.04 | 0.72-1.50 | 0.84 |
| **Fasting sample (n=3,576)** |  |  |  |  |  |  |
| Triglycerides (mg/dL) | 1.34 | 1.27-1.41 | <0.001 | 1.11 | 1.07-1.16 | <0.001 |
| Diabetes |  |  |  |  |  |  |
| Normal glucose | 1.0 |  |  | 1.0 |  |  |
| Pre-diabetes | 2.52 | 1.90-3.36 | <0.001 | 2.06 | 0.97-4.39 | 0.059 |
| Diabetes | 9.48 | 6.30-14.3 | <0.001 | 8.09 | 4.15-15.76 | <0.001 |
| Insulin (pmol/L) | 1.54 | 1.49-1.60 | <0.001 | 1.29 | 1.17-1.41 | <0.001 |

OR, odds ratio; CI, confidence interval; CAP, controlled attenuation parameter; BMI, body mass index; HDL, high-density lipoprotein; ALT, alanine aminotransferase; AST, aspartate aminotransferase; GGT, gamma-glutamyltransferase; AA, Associate in Arts.
<sup>1</sup>Models for age were unadjusted.
<sup>2</sup>Moderate alcohol use was <3 drinks/day for men and <2 drinks/day for women; heavy alcohol use was $\geq 3$ drinks/day for men and $\geq 2$ drinks/day for women.
<sup>3</sup>Does not meet physical activity guidelines.

**Table 4.** Multivariate-adjusted logistic regression associations of fatty liver disease and fibrosis risk among participants 20+ years in the National Health and Nutrition Examination Survey, 2017-March 2020 prepandemic data

| Characteristic | Fatty liver disease<br>(>300 dB/m) (N=6,625) |  |  | Fibrosis risk<br>(>8 kPa) (N=6,776) |  |  |
| --- | --- | --- | --- | --- | --- | --- |
|  | OR | 95 % CI | p-value | OR | 95 % CI | p-value |
| Age (years) | 1.01 | 1.00-1.01 | 0.017 | 1.01 | 1.00-1.02 | 0.12 |
| Sex |  |  |  |  |  |  |
| Men | 1.0 |  |  | 1.0 |  |  |
| Women | 1.12 | 0.91-1.37 | 0.27 | 0.89 | 0.59-1.36 | 0.59 |
| Race-ethnicity |  |  |  |  |  |  |
| Non-Hispanic white | 1.0 |  |  | 1.0 |  |  |
| Non-Hispanic black | 0.51 | 0.39-0.65 | <0.001 | 0.78 | 0.53-1.15 | 0.20 |
| Non-Hispanic Asian | 1.49 | 1.05-2.10 | 0.027 | 1.21 | 0.77-1.90 | 0.40 |
| Hispanic | 1.17 | 0.86-1.59 | 0.31 | 0.94 | 0.65-1.35 | 0.72 |
| BMI (kg/m<sup>2</sup>) | 1.14 | 1.12-1.16 | <0.001 | 1.11 | 1.09-1.13 | <0.001 |
| Waist-to-hip ratio | 1.07 | 1.05-1.08 | <0.001 | 1.02 | 1.00-1.04 | 0.064 |
| Diabetes |  |  |  |  |  |  |
| Normal glucose | 1.0 |  |  | 1.0 |  |  |
| Pre-diabetes | 1.47 | 1.15-1.87 | 0.004 | 1.24 | 0.92-1.68 | 0.15 |
| Diabetes | 2.58 | 1.90-3.51 | <0.001 | 2.56 | 1.93-3.38 | <0.001 |
| Total cholesterol deciles | 1.07 | 1.04-1.10 | <0.001 | 0.95 | 0.92-0.98 | 0.006 |
| HDL cholesterol deciles | 0.89 | 0.84-0.95 | <0.001 | -- | -- | -- |
| Blood pressure elevated | 1.28 | 1.03-1.59 | 0.030 | 1.73 | 1.36-2.20 | <0.001 |
| C-reactive protein deciles | 1.06 | 1.01-1.11 | 0.014 | -- | -- | -- |
| ALT deciles | 1.20 | 1.14-1.26 | <0.001 | -- | -- | -- |
| AST deciles | -- | -- | -- | 1.17 | 1.11-1.24 | <0.001 |
| CAP deciles | -- | -- | -- | 1.07 | 1.02-1.13 | 0.012 |
| Physical inactivity<sup>1</sup> | 1.26 | 0.99-1.59 | 0.056 | -- | -- | -- |
| Sedentary behavior ≥ 12 hours/day | 1.36 | 0.96-1.91 | 0.077 | -- | -- | -- |
| Diet healthiness |  |  |  |  |  |  |
| Excellent / very good | 1.0 |  |  | 1.0 |  |  |
| Good | 1.14 | 0.86-1.52 | 0.35 | 1.18 | 0.84-1.65 | 0.34 |
| Fair / poor | 1.28 | 0.97-1.69 | 0.078 | 1.43 | 1.00-2.05 | 0.048 |
| Education |  |  |  |  |  |  |
| Below high school graduate | -- | -- | -- | 1.29 | 0.85-1.96 | 0.22 |
| High school graduate | -- | -- | -- | 1.77 | 1.05-3.00 | 0.034 |
| Some college or AA degree | -- | -- | -- | 1.28 | 0.90-1.82 | 0.16 |
| College graduate or above | -- | -- | -- | 1.0 |  |  |

OR, odds ratio; CI, confidence interval; BMI, body mass index; HDL, high-density lipoprotein; ALT, alanine aminotransferase; AST, aspartate aminotransferase; CAP, controlled attenuation parameter; AA, Associate in Arts.
<sup>1</sup>Does not meet physical activity guidelines.

**Table 5.** Multivariate-adjusted logistic regression associations of fatty liver disease and fibrosis risk among participants 20+ years in the National Health and Nutrition Examination Survey, 2017-March 2020 prepandemic data **(fasting sample, N=3**,**576)**

| Characteristic | Fatty liver disease<br>(>300 dB/m) (N=3,351) |  |  | Fibrosis risk<br>(>8 kPa) (N=2,830) |  |  |
| --- | --- | --- | --- | --- | --- | --- |
|  | OR | 95 % CI | p-value | OR | 95 % CI | p-value |
| Age (years) | 1.01 | 1.00-1.02 | 0.091 | 1.01 | 0.99-1.03 | 0.17 |
| Sex |  |  |  |  |  |  |
| Men | 1.0 |  |  | 1.0 |  |  |
| Women | 0.87 | 0.65-1.16 | 0.33 | 0.95 | 0.58-1.55 | 0.84 |
| Race-ethnicity |  |  |  |  |  |  |
| Non-Hispanic white | 1.0 |  |  | 1.0 |  |  |
| Non-Hispanic black | 0.47 | 0.30-0.74 | 0.002 | 0.50 | 0.22-1.17 | 0.11 |
| Non-Hispanic Asian | 1.39 | 1.00-1.94 | 0.049 | 1.42 | 0.72-2.79 | 0.29 |
| Hispanic | 1.01 | 0.73-1.40 | 0.94 | 1.01 | 0.58-1.77 | 0.96 |
| BMI (kg/m<sup>2</sup>) | 1.14 | 1.11-1.18 | <0.001 | 1.10 | 1.06-1.14 | <0.001 |
| Waist-to-hip ratio | 1.06 | 1.04-1.09 | <0.001 | -- | -- | -- |
| Diabetes |  |  |  |  |  |  |
| Normal glucose | 1.0 |  |  | 1.0 |  |  |
| Pre-diabetes | 0.93 | 0.58-1.47 | 0.74 | 0.97 | 0.47-1.99 | 0.93 |
| Diabetes | 2.13 | 1.26-3.62 | 0.007 | 2.17 | 1.12-4.21 | 0.024 |
| Triglyceride deciles | 1.10 | 1.04-1.17 | 0.003 | -- | -- | -- |
| Insulin deciles | 1.18 | 1.12-1.25 | <0.001 | -- | -- | -- |
| Total cholesterol deciles | 1.08 | 1.03-1.14 | 0.004 | 0.94 | 0.89-0.99 | 0.022 |
| Blood pressure elevated | -- | -- | -- | 1.73 | 1.13-2.65 | 0.013 |
| ALT deciles | 1.08 | 1.00-1.17 | 0.039 |  |  |  |
| GGT deciles | -- | -- | -- | 1.20 | 1.10-1.32 | <0.001 |
| Sedentary behavior ≥ 12 hours/day | 2.29 | 1.30-4.03 | 0.006 | 1.68 | 1.10-2.56 | 0.019 |
| Poverty to income ratio |  |  |  |  |  |  |
| <1.65 | -- | -- | -- | 2.00 | 1.18-3.40 | 0.012 |
| 1.65 - <3.20 | -- | -- | -- | 2.59 | 1.38-4.85 | 0.005 |
| 3.20 - <5 | -- | -- | -- | 1.16 | 0.48-2.82 | 0.73 |
| ≥5 | -- | -- | -- | 1.0 |  |  |

OR, odds ratio; CI, confidence interval; BMI, body mass index; ALT, alanine aminotransferase; GGT, gamma-glutamyltransferase.

### Fibrosis risk

Participants with fibrosis were less likely to be non-Hispanic Asian **(Table 2)**. When examined by sex, this was primarily due to lower fibrosis prevalence among non-Hispanic Asian women (**Figure 1**). Among persons with fibrosis, only 10.9% reported ever having been told by a health care provider that they had liver disease and only 8.1% reported still having liver disease. Persons with fibrosis were older and more likely to have metabolic risk factors, higher liver enzymes, and to be former smokers, and less likely to be moderate drinkers. They were less likely to meet physical activity guidelines, more likely to be sedentary for 12 or more hours a day, and self-reported a less healthy diet **(Table 2, Figure 2a)**. Fibrosis prevalence was highest among persons who did not meet physical activity guidelines and were sedentary for 12 or more hours a day **(Figure 2b)**. The higher fibrosis prevalence among persons with less favorable lifestyle habits was seen across sex and race-ethnicity groups, although differences did not reach statistical significance among all groups (**Supplemental Figures 2-6)**. Participants with fibrosis were less likely to have a college degree and had lower income **(Table 2, Supplemental Figure 7)**. The lower fibrosis prevalence among persons with the highest education level was seen across sex and race-ethnicity groups, although differences did not reach statistical significance among all groups (**Supplemental Figures 8-11)**. ALT, AST, and GGT were elevated among only 28.4%, 19.7%, and 34.2% of persons with fibrosis, respectively **(Table 2)**.

With adjustment for age, there was no longer a relationship with alcohol use (**Table 3**). In multivariate-adjusted analysis, fibrosis remained associated with diabetes, higher BMI, blood pressure, AST, and CAP, and inversely with total cholesterol. There were also associations with a less healthy diet and less education (**Table 4**). Among persons who fasted, factors most strongly associated with fibrosis in multivariate-adjusted analysis were diabetes, higher BMI, blood pressure, GGT, and sedentary time, and lower total cholesterol and income (**Table 5**). Because of the smaller sample size compared with **Table 4**, some factors from the full sample model no longer remained associated (i.e. waist-to-hip ratio, CAP, and diet) and income was more strongly related than education. In contrast, greater sedentary behavior became associated with fibrosis.

## DISCUSSION

In this representative U.S. population sample with transient elastography, the prevalence of fatty liver disease using a CAP cut point of >300 dB/m was high (28.8%) and requires scalable management approaches at a population level.^5-6^ The prevalence of fibrosis, defined as liver stiffness >8 kPa, was also high overall (10.4%), especially among persons with fatty liver disease (22.8%). Fatty liver disease prevalence estimates ranged from 35.1% using a cut point of 285 dB/m, to 47.8% with a cut point of 263 dB/m, to over half of the U.S. population (56.7%) using a cut point of 248 dB/m in analyses of NHANES 2017-2018.^18-19^ Prevalence of hepatic steatosis and fibrosis was even higher among some U.S. population groups, such as persons with type 2 diabetes mellitus.^30^ The fibrosis prevalence estimated in the general U.S. population was higher compared with a Dutch population study that used the same liver stiffness cut point of 8.0 kPa (10.4% vs. 6.0%), despite the older age of the Dutch population sample (45 years and over).^9^ Among European population-based studies using transient elastography for screening with cut points ranging from 7.9 kPa to 9.6 kPa, fibrosis prevalence varied from 2.4% to 7.5%.^31-36^ Among Asian population studies with cut points ranging from 5.9 kPa to 9.6 kPa, fibrosis prevalence varied from 2.0% to 14.3%.^31,37-39^ These reports were limited to community-based samples or persons undergoing health examinations with smaller samples than NHANES 2017-March 2020.

The proposed MAFLD definition had limited effect on prevalence estimates in the U.S. population compared with the NAFLD definition (28% vs. 26%). The prevalence of fibrosis was similar for the two conditions (23% for each). MAFLD and NAFLD prevalence were also similar, 39.1% vs. 37.1%, in an analysis of NHANES 2017-2018 using 274 dB/m to identify liver steatosis.^8^ A prospective population study from Hong Kong using proton-magnetic resonance spectroscopy found a similar prevalence of MAFLD and NAFLD (25.9% vs. 25.7%), but reported a lower incidence of MAFLD compared with NAFLD, especially among persons with low BMI.^10^ A Dutch population-based study found MAFLD and NAFLD prevalence of 34.3% and 29.8%, respectively, using ultrasound and persons with MAFLD only, but not those with NAFLD only, were more likely to have fibrosis.^9^ MAFLD was found to be more strongly associated with all-cause mortality compared with NAFLD in the U.S. population.^40^ However, additional longitudinal studies of the long-term effects of the proposed definition will be needed.

The extent of undiagnosed liver disease in the U.S. population is striking and requires a unified public health response including risk stratification by primary care providers.^5^ Among persons with fatty liver disease, only 7% reported ever having been told by a health care provider that they had liver disease and among persons with more severe disease represented by fibrosis, only a minimally higher proportion, 11%, reported ever having been told they had liver disease. Liver disease awareness has improved little from previous reports in the U.S. population using earlier NHANES data in which NAFLD was identified by liver fat scores.^41^ Fatty liver disease may remain asymptomatic and not come to expert medical attention until reaching an advanced stage. However, despite the current lack of an approved pharmacologic therapy, persons identified early with fatty liver disease can benefit from recommendations for diet and exercise for weight loss, as well as treatment of cardiovascular disease risk factors. The low liver disease awareness that we report indicates that even among high-risk groups, many persons are not being evaluated for fatty liver disease despite the availability of noninvasive screening techniques such as transient elastography and liver fat and fibrosis scores. Routine screening for NAFLD in high-risk groups attending primary care, diabetes, or obesity clinics is not currently recommended by the American Association for the Study of Liver Diseases.^27^ However, these are often the health care settings in which patients at high risk for fatty liver disease are first seen. Currently, there is insufficient awareness of the growing NAFLD burden among both the public and the health care providers caring for them. A recent global survey of physicians’ knowledge about NAFLD found a significant knowledge gap for the identification, diagnosis, and management of NAFLD, especially among primary care providers who are often the first to see patients at risk for NAFLD.^42^ To address this challenge, a Clinical Care Pathway has recently been developed by the American Gastroenterological Association to provide guidance on the screening, diagnosis, and treatment of NAFLD.^6^

We found associations with known fatty liver disease risk factors, including age, non-Hispanic Asian race-ethnicity, BMI, waist-to-hip ratio, pre-diabetes and diabetes, total cholesterol, blood pressure, C-reactive protein and ALT, and inverse relationships with non-Hispanic black race-ethnicity and HDL cholesterol. Fibrosis was associated with BMI, diabetes, blood pressure, AST, CAP, and total cholesterol (inverse). Lower cholesterol among persons with fibrosis was previously reported using earlier NHANES data.^13,43^ In the current analysis we also examined lifestyle factors and found that persons with fatty liver disease were less likely to meet physical activity guidelines, more likely to be sedentary for 12 or more hours a day, and self-reported a less healthy diet. Fatty liver disease prevalence was highest among persons with both a lower level of physically activity and a higher level of sedentary behavior. Associations with physical activity, sedentary behavior, and diet were only partially explained after accounting for demographic, clinical, and other lifestyle factors. Similar associations were seen for fibrosis, though relationships with physical activity and sedentary behavior did not remain after accounting for other factors, possibly due to heterogeneity and smaller numbers of persons with fibrosis compared with fatty liver disease. Further analysis of persons who fasted revealed a stronger association of sedentary behavior with fatty liver disease and fibrosis suggesting that accounting for triglycerides and/or insulin may have augmented this relationship similar to an earlier CDC report that sedentary behavior was associated with higher insulin levels in the U.S. population.^44^ The higher fatty liver disease and fibrosis prevalence among persons with detrimental lifestyle habits was seen across sex and race-ethnicity groups, although differences did not reach statistical significance among all groups due to heterogeneity and smaller sample sizes. These findings suggest that adoption of beneficial physical activity and dietary habits could have broad impact across U.S. population groups on the fatty liver disease burden. Beneficial effects of a healthier lifestyle on fatty liver disease and fibrosis were also reported in previous papers using NHANES 2017-2018 data.^12-14^ Despite the potential benefit of lifestyle interventions across demographic groups, the COVID-19 pandemic has had a detrimental effect on lifestyle habits of less advantaged groups. A recent systematic review found decreased physical activity levels during the COVID-19 lockdown across almost all reviewed populations and increased sedentary behavior in the majority of studies included.^45^ If these trends continue, they could exacerbate the growing burden of fatty liver disease and fibrosis in the U.S. population.

We examined the relationship of socioeconomic status with fatty liver disease and fibrosis using educational attainment and poverty income ratio (ratio of family income to poverty threshold). As an example of poverty income ratio, the poverty threshold was $26,200 for a family of 4 in 2020.^46^ Therefore, a family of 4 with a total income of $26,200 would have a poverty income ratio of 1.0, while a family of 4 with a total income of $43,230, $83,840, or $131,000 would have a poverty-income ratio of 1.65, 3.20, or 5.0, respectively, the cut points used in this analysis. Persons with a college degree or higher and those with the highest income (poverty income ratio ≥5) were less likely to have fatty liver disease and fibrosis, though the relationship of income with fatty liver disease was not statistically significant. Further analysis of persons who fasted revealed a stronger association of income with fibrosis.

A limitation of the use of transient elastography is the lack of universally accepted cut points for identifying hepatic steatosis and fibrosis in the general population. This limits the ability to compare fatty liver disease and fibrosis prevalence and associations across studies. We used previously reported and validated cut points for our analysis.^26^ Another limitation was the sole use of self-reported diet healthiness as a measure of diet quality because 24-hour dietary recall data have not yet been released for the NHANES 2017-March 2020 prepandemic survey cycle. However, we found a strong association between a self-reported less healthy diet and higher prevalence of fatty liver disease and fibrosis. These relationships should be explored further when 24-hour dietary recall data become available. Similarly, we were unable to evaluate the relationship of viral hepatitis with CAP and liver stiffness or to exclude participants with viral hepatitis in identifying NAFLD due to the lack of data on viral hepatitis B and C serum markers that were measured, but have not yet been released. However, among the NHANES 2017-2018 sample, only 0.28% were hepatitis B virus positive and only 0.99% and 1.0%, respectively, were RNA negative, antibody positive and RNA positive for hepatitis C virus.^21^ Finally, the cross-sectional design using survey data did not allow determination of causation. Despite limitations, NHANES 2017-March 2020 provides the only U.S. nationally representative transient elastography biomarker data. Strengths include avoidance of ascertainment bias found in clinical studies of conveniently selected patients and ability to generalize results to U.S. population ethnic-racial groups that are differentially represented in most studies. Other benefits include the well-characterized sample and wealth of data available on potential fatty liver disease risk factors including socioeconomic status.

In conclusion, fatty liver disease and fibrosis prevalence are high in the U.S. population and most persons are unaware of having liver disease. Fatty liver disease can progress to severe fibrosis with increased risk of liver-related complications and death.^3,4^ Consequently, a multi-pronged approach toward a unified public health response is needed, including risk stratification by primary care providers. Given that changes in lifestyle factors might positively influence the fatty liver disease burden, there is an urgent need for screening and fatty liver disease management in high-risk individuals using transient elastography or other noninvasive methods to intervene in disease progression.

## Supporting information

Strobe checklist

## Data Availability

All NHANES data used for this paper are publicly available.

https://www.cdc.gov/nchs/nhanes/index.htm

## ACKNOWLEDGMENTS

The National Institute of Diabetes and Digestive and Kidney Diseases is a funding collaborator with the Centers for Disease Control and Prevention (CDC) National Center for Health Statistics (NCHS) through the United States Government Interagency Agreement (IAA) Number 15006 (also known as IAA# 164 at NCHS).

Author names in bold designate shared co-first authorship.

## REFERENCES

1. Peery AF, Crockett SD, Murphy CC, et al. Burden and Cost of Gastrointestinal, Liver, and Pancreatic Diseases in the United States: Update 2021. Gastroenterology 2022;162:621–644.

2. Xu JQ, Murphy SL, Kochanek KD, et al. Deaths: Final data for 2019. National Vital Statistics Reports; vol 70 no 08. Hyattsville, MD: National Center for Health Statistics. 2021.

3. Unalp-Arida A, Ruhl CE. Noninvasive fatty liver markers predict liver disease mortality in the U.S. population. Hepatology 2016;63:1170–1183.

4. Sanyal AJ, Van Natta ML, Clark J, et al.; NASH Clinical Research Network (CRN). Prospective Study of Outcomes in Adults with Nonalcoholic Fatty Liver Disease. N Engl J Med 2021;385:1559–1569.

5. Kanwal F, Shubrook JH, Younossi Z, et al. Preparing for the NASH Epidemic: A Call to Action. Gastroenterology 2021;161:1030–1042.

6. Kanwal F, Shubrook JH, Adams LA, et al. Clinical Care Pathway for the Risk Stratification and Management of Patients With Nonalcoholic Fatty Liver Disease. Gastroenterology 2021;161:1657–1669.

7. Eslam M, Newsome PN, Sarin SK, et al. A new definition for metabolic dysfunction-associated fatty liver disease: An international expert consensus statement. J Hepatol 2020;73:202–209.

8. Ciardullo S, Perseghin G. Prevalence of NAFLD, MAFLD and associated advanced fibrosis in the contemporary United States population. Liver Int 2021;41:1290–1293.

9. van Kleef LA, Ayada I, Alferink LJM, et al. Metabolic dysfunction-associated fatty liver disease improves detection of high liver stiffness: The Rotterdam Study. Hepatology 2022;75:419–429.

10. Wong VW, Wong GL, Woo J, et al. Impact of the New Definition of Metabolic Associated Fatty Liver Disease on the Epidemiology of the Disease. Clin Gastroenterol Hepatol 2021;19:2161–2171.

11. Younossi ZM, Corey KE, Lim JK. AGA Clinical Practice Update on Lifestyle Modification Using Diet and Exercise to Achieve Weight Loss in the Management of Nonalcoholic Fatty Liver Disease: Expert Review. Gastroenterology 2021;160:912–918.

12. Kim D, Konyn P, Cholankeril G, et al. Physical Activity Is Associated With Nonalcoholic Fatty Liver Disease and Significant Fibrosis Measured by FibroScan. Clin Gastroenterol Hepatol 2021 Jun 29:S1542-3565(21)00693-5.

13. Vilar-Gomez E, Nephew LD, Vuppalanchi R, et al. High-quality diet, physical activity, and college education are associated with low risk of NAFLD among the US population. Hepatology 2021 Oct 20. doi: 10.1002/hep.32207.

14. Heredia NI, Zhang X, Balakrishnan M, et al. Physical activity and diet quality in relation to non-alcoholic fatty liver disease: A cross-sectional study in a representative sample of U.S. adults using NHANES 2017-2018. Prev Med 2022 Jan;154:106903.

15. National Institute of Diabetes and Digestive and Kidney Diseases. NIDDK Strategic Plan for Research. Dec. 2021. Internet: https://www.niddk.nih.gov/about-niddk/strategic-plans-reports/niddk-strategic-plan-for-research (accessed February, 2022).

16. Sasso M, Miette V, Sandrin L, et al. The controlled attenuation parameter (CAP): A novel tool for the non-invasive evaluation of steatosis using Fibroscan^®^. Clin Res Hepatol Gastroenterol 2012;36:13–20.

17. Fabrellas N, Hernandez R, Graupera I, et al. Prevalence of hepatic steatosis as assessed by controlled attenuation parameter (CAP) in subjects with metabolic risk factors in primary care. A population-based study. PLoS ONE 2018;13:e0200656.

18. Kim D, Cholankeril G, Loomba R, et al. Prevalence of Fatty Liver Disease and Fibrosis Detected by Transient Elastography in Adults in the United States, 2017-2018. Clin Gastroenterol Hepatol 2021;19:1499–1501.

19. Zhang X, Heredia NI, Balakrishnan M, et al. Prevalence and factors associated with NAFLD detected by vibration controlled transient elastography among US adults: Results from NHANES 2017-2018. PLoS One 2021 Jun 3;16(6):e0252164.

20. National Center for Health Statistics. National Health and Nutrition Examination Survey 2017-2018 Liver Ultrasound Transient Elastography Procedures Manual. Internet: https://www.n.cdc.gov/nchs/data/nhanes/2017-2018/manuals/2018_Liver_Ultrasound_Elastography_Procedures_Manual.pdf (accessed February 2022).

21. Unalp-Arida A, Ruhl CE. Transient Elastography-Assessed Hepatic Steatosis and Fibrosis Are Associated With Body Composition in the United States. Clin Gastroenterol Hepatol 2021 Feb 5:S1542-3565(21)00110-5.

22. National Center for Health Statistics. National Health and Nutrition Examination Survey. Internet: https://www.cdc.gov/nchs/nhanes/index.htm (accessed February 2022).

23. World Health Organization. Global Physical Activity Questionnaire (GPAQ) Analysis Guide. Internet: https://www.who.int/ncds/surveillance/steps/resources/GPAQ_Analysis_Guide.pdf (accessed February 2022).

24. Vuppalanchi R, Siddiqui MS, Van Natta ML, et al.; NASH Clinical Research Network. Performance characteristics of vibration-controlled transient elastography for evaluation of nonalcoholic fatty liver disease. Hepatology 2018;67:134–144.

25. Siddiqui MS, Vuppalanchi R, Van Natta ML, et al.; NASH Clinical Research Network.Vibration-Controlled Transient Elastography to Assess Fibrosis and Steatosis in Patients With Nonalcoholic Fatty Liver Disease. Clin Gastroenterol Hepatol 2019;17:156–163.

26. Eddowes PJ, Sasso M, Allison M, et al. Accuracy of FibroScan controlled attenuation parameter and liver stiffness measurement in assessing steatosis and fibrosis in patients with nonalcoholic fatty liver disease. Gastroenterology 2019;156:1717–1730.

27. Chalasani N, Younossi Z, Lavine JE, et al. The diagnosis and management of nonalcoholic fatty liver disease: Practice guidance from the American Association for the Study of Liver Diseases. Hepatology 2018;67:328–357.

28. Hosmer DW, Lemeshow S, Sturdivant RX. Applied Logistic Regression. 3rd ed. Hoboken, NJ: John Wiley & Sons, Inc., 2013:89–149.

29. Breslow NE, Day NE. Statistical Methods in Cancer Research: the Design and Analysis of Cohort Studies. Lyon, France: International Agency for Research on Cancer, 1987:48–79.

30. Ciardullo S, Monti T, Perseghin G. High Prevalence of Advanced Liver Fibrosis Assessed by Transient Elastography Among U.S. Adults With Type 2 Diabetes. Diabetes Care 2021;44:519–525.

31. Ginès P, Castera L, Lammert F, et al.; LiverScreen Consortium Investigators. Population screening for liver fibrosis: Toward early diagnosis and intervention for chronic liver diseases. Hepatology 2022;75:219–228.

32. Roulot D, Costes JL, Buyck JF, et al. Transient elastography as a screening tool for liver fibrosis and cirrhosis in a community-based population aged over 45 years. Gut 2011;60:977–984.

33. Koehler EM, Plompen EP, Schouten JN, et al. Presence of diabetes mellitus and steatosis is associated with liver stiffness in a general population: The Rotterdam study. Hepatology 2016;63:138–147.

34. Caballería L, Pera G, Arteaga I, et al. High Prevalence of Liver Fibrosis Among European Adults With Unknown Liver Disease: A Population-Based Study. Clin Gastroenterol Hepatol 2018;16:1138–1145.

35. Petta S, Di Marco V, Pipitone RM, et al. Prevalence and severity of nonalcoholic fatty liver disease by transient elastography: Genetic and metabolic risk factors in a general population. Liver Int 2018;38:2060–2068.

36. Abeysekera KWM, Fernandes GS, Hammerton G, et al. Prevalence of steatosis and fibrosis in young adults in the UK: a population-based study. Lancet Gastroenterol Hepatol 2020;5:295–305.

37. Wong VW, Chu WC, Wong GL, et al. Prevalence of non-alcoholic fatty liver disease and advanced fibrosis in Hong Kong Chinese: a population study using proton-magnetic resonance spectroscopy and transient elastography. Gut 2012;61:409–415.

38. Baba M, Furuya K, Bandou H, et al. Discrimination of individuals in a general population at high-risk for alcoholic and non-alcoholic fatty liver disease based on liver stiffness: a cross section study. BMC Gastroenterol 2011 Jun 13;11:70.

39. You SC, Kim KJ, Kim SU, et al. Factors associated with significant liver fibrosis assessed using transient elastography in general population. World J Gastroenterol 2015;21:1158–1166.

40. Kim D, Konyn P, Sandhu KK, et al. Metabolic dysfunction-associated fatty liver disease is associated with increased all-cause mortality in the United States. J Hepatol 2021;75:1284–1291.

41. Alqahtani SA, Paik JM, Biswas R, et al. Poor Awareness of Liver Disease Among Adults With NAFLD in the United States. Hepatol Commun 2021;5:1833–1847.

42. Younossi ZM, Ong JP, Takahashi H, et al.; Global Nonalcoholic Steatohepatitis Council. A Global Survey of Physicians Knowledge About Nonalcoholic Fatty Liver Disease. Clin Gastroenterol Hepatol 2021 Jul 3:S1542-3565(21)00719-9.

43. Unalp-Arida A, Ruhl CE. Liver fibrosis scores predict liver disease mortality in the United States population. Hepatology 2017;66:84–95.

44. Ford ES, Li C, Zhao G, et al. Sedentary behavior, physical activity, and concentrations of insulin among US adults. Metabolism 2010;59:1268–1275.

45. Stockwell S, Trott M, Tully M, et al. Changes in physical activity and sedentary behaviours from before to during the COVID-19 pandemic lockdown: a systematic review. BMJ Open Sport Exerc Med 2021 Feb 1;7(1):e000960.

46. Office of the Assistant Secretary for Planning and Evaluation. 2020 Poverty Guidelines. Internet: https://aspe.hhs.gov/topics/poverty-economic-mobility/poverty-guidelines/prior-hhs-poverty-guidelines-federal-register-references/2020-poverty-guidelines (accessed February 2022).

