## Supplemental tables and figure legends for "Prepandemic prevalence estimates of fatty liver disease and fibrosis defined by liver elastography in the United States"

**Supplemental Table 1.** Characteristics of participants 20+ years with complete and partial liver elastography in the National Health and Nutrition Examination Survey, United States, 2017-March 2020

| Characteristic^1^ | Transient elastography | | | |
| --- | --- | --- | --- | --- |
|  | Complete  (n=7,396) | Partial | | |
|  |  | Fasted <3 hours  (n=248) | 1 to <10 valid measures  (n=99) | Stiffness IQR/median ≥30%  (n=180) |
| Women | 50.6 | 52.2 | 56.2 | 53.9 |
| Race-ethnicity |  |  |  |  |
| Non-Hispanic white | 62.7 | 59.4 | 62.7 | 73.2^2^ |
| Non-Hispanic black | 11.2 | 12.6 | 12.8 | 10.8 |
| Non-Hispanic Asian | 5.9 | 7.1 | 4.9 | 2.8^2^ |
| Hispanic | 16.1 | 18.4 | 15.2 | 9.5^2^ |
| Age (years) | 48.1 (17.2) | 45.0 (17.5) | 57.0 (14.9)^2^ | 52.9 (16.5)^2^ |
| BMI (kg/m^2^) | 29.5 (7.0) | 28.2 (7.8)^2^ | 35.8 (11.6)^2^ | 37.7 (9.8)^2^ |
| ALT (IU/L) | 23.0 (17.5) | 19.0 (12.5)^2^ | 22.4 (23.5) | 23.1 (14.2) |
| AST (IU/L) | 21.9 (12.8) | 21.1 (10.5) | 20.4 (9.0) | 22.3 (15.1) |
| GGT (IU/L) | 29.5 (37.9) | 23.5 (26.2)^2^ | 26.7 (21.6) | 42.7 (152.8) |
| Number of complete measures^3^ | 11.2 (2.2) | 11.3 (2.0) | 5.2 (2.9)^2^ | 11.1 (3.4) |
| Number of attempted measures^3^ | 14.2 (5.2) | 13.9 (4.6) | 26.6 (5.9)^2^ | 23.1 (7.8)^2^ |
| CAP (dB/m) |  |  |  |  |
| Median | 263.9 (62.4) | 244.4 (58.4)^2^ | 310.6 (70.8)^2^ | 302.5 (72.2)^2^ |
| IQR | 36.8 (19.8) | 40.4 (26.2) | 28.4 (21.5)^2^ | 39.3 (25.1) |
| Liver stiffness (kPa) |  |  |  |  |
| Median | 5.7 (4.4) | 6.1 (6.9) | 11.9 (12.9)^2^ | 10.4 (8.6)^2^ |
| IQR | 0.8 (0.8) | 0.9 (0.9) | 4.3 (8.4)^2^ | 7.2 (8.5)^2^ |
| IQR/median (%) | 13.8 (6.1) | 14.2 (6.4) | 44.4 (108.9)^2^ | 67.7 (62.3)^2^ |

IQR, interquartile range; BMI, body mass index; ALT, alanine aminotransferase; AST, aspartate aminotransferase; GGT, gamma-glutamyltransferase; CAP, controlled attenuation parameter.

^1^Values are % or mean (standard deviation).

^2^p<0.05 compared with complete exam.

^3^Elastography measures using the final probe.

**Supplemental Table 2.** Decile cut points for the 10^th^ to 90^th^ percentiles of clinical variables among participants 20+ years in the National Health and Nutrition Examination Survey, 2017-March 2020 prepandemic data

|  | Decile | | | | | | | | |
| --- | --- | --- | --- | --- | --- | --- | --- | --- | --- |
|  | 10th | 20th | 30th | 40th | 50th | 60th | 70th | 80th | 90th |
| Full sample (n=7,923) | | | | | | | | | |
| Total cholesterol (mg/dL) | 138.0 | 152.2 | 163.5 | 173.6 | 183.3 | 193.7 | 205.2 | 218.8 | 239.1 |
| HDL cholesterol (mg/dL) | 35.8 | 40.3 | 43.6 | 47.4 | 50.7 | 54.6 | 59.3 | 65.2 | 73.6 |
| CRP (mg/L) | 0.4 | 0.7 | 1.0 | 1.3 | 1.8 | 2.6 | 3.6 | 5.2 | 8.2 |
| ALT (IU/L) | 9.8 | 12.0 | 13.6 | 15.5 | 17.5 | 20.1 | 23.5 | 28.8 | 38.2 |
| AST (IU/L) | 13.3 | 14.8 | 16.1 | 17.4 | 18.7 | 20.0 | 22.0 | 24.6 | 29.7 |
| GGT (IU/L) | 9.8 | 12.3 | 14.3 | 16.8 | 19.5 | 22.9 | 27.0 | 34.5 | 51.2 |
| CAP (dB/m) | 186.7 | 209.0 | 225.0 | 242.4 | 261.5 | 278.8 | 297.5 | 319.5 | 352.1 |
| Fasting sample (n=3,576) | | | | | | | | | |
| Triglycerides (mg/dL) | 43.7 | 54.8 | 65.6 | 77.0 | 88.7 | 104.6 | 122.3 | 148.6 | 192.3 |
| Insulin (pmol/L) | 24.0 | 31.4 | 38.9 | 46.7 | 56.6 | 68.5 | 82.8 | 106.8 | 151.5 |

HDL, high-density lipoprotein; CRP, C-reactive protein; ALT, alanine aminotransferase; AST, aspartate aminotransferase; GGT, gamma-glutamyltransferase; CAP, controlled attenuation parameter.

**SUPPLEMENTAL FIGURE LEGENDS**

Supplemental Figure 1. Analysis sample for NHANES 2017-March 2020 prepandemic transient elastography data

NHANES, National Health and Nutrition Examination Survey.

Supplemental Figure 2. Prevalence of fatty liver disease and fibrosis by **physical activity or sedentary time** among **men and women** 20+ years in the National Health and Nutrition Examination Survey, United States, 2017-March 2020 prepandemic data (N=7,923)

Supplemental Figure 3. Prevalence of fatty liver disease and fibrosis by **diet healthiness** and **sex** among participants 20+ years in the National Health and Nutrition Examination Survey, United States, 2017-March 2020 prepandemic data (N=7,923)

Supplemental Figure 4. Prevalence of fatty liver disease and fibrosis by **physical activity** and **race-ethnicity** among participants 20+ years in the National Health and Nutrition Examination Survey, United States, 2017-March 2020 prepandemic data (N=7,923)

Supplemental Figure 5. Prevalence of fatty liver disease and fibrosis by **sedentary time** and **race-ethnicity** among participants 20+ years in the National Health and Nutrition Examination Survey, United States, 2017-March 2020 prepandemic data (N=7,923)

Supplemental Figure 6. Prevalence of fatty liver disease and fibrosis by **diet healthiness** and **race-ethnicity** among participants 20+ years in the National Health and Nutrition Examination Survey, United States, 2017-March 2020 prepandemic data (N=7,923)

Supplemental Figure 7. Prevalence of fatty liver disease and fibrosis by **social factors** among participants 20+ years in the National Health and Nutrition Examination Survey, United States, 2017-March 2020 prepandemic data (N=7,923)

Supplemental Figure 8. Prevalence of fatty liver disease and fibrosis by **education** and **sex** among participants 20+ years in the National Health and Nutrition Examination Survey, United States, 2017-March 2020 prepandemic data (N=7,923)

Supplemental Figure 9. Prevalence of fatty liver disease and fibrosis by **poverty income ratio** and **sex** among participants 20+ years in the National Health and Nutrition Examination Survey, United States, 2017-March 2020 prepandemic data (N=7,923)

Supplemental Figure 10. Prevalence of fatty liver disease and fibrosis by **education** and **race-ethnicity** among participants 20+ years in the National Health and Nutrition Examination Survey, United States, 2017-March 2020 prepandemic data (N=7,923) **(A).** Non-Hispanic whites and non-Hispanic blacks. **(B).** Non-Hispanic Asians and Hispanics.

Supplemental Figure 11. Prevalence of fatty liver disease and fibrosis by **poverty income ratio** and **race-ethnicity** among participants 20+ years in the National Health and Nutrition Examination Survey, United States, 2017-March 2020 prepandemic data (N=7,923) **(A).** Non-Hispanic whites and non-Hispanic blacks. **(B).** Non-Hispanic Asians and Hispanics.
