## Supplementary figures and images for "Prepandemic prevalence estimates of fatty liver disease and fibrosis defined by liver elastography in the United States"

### Supplemental Figure 1.

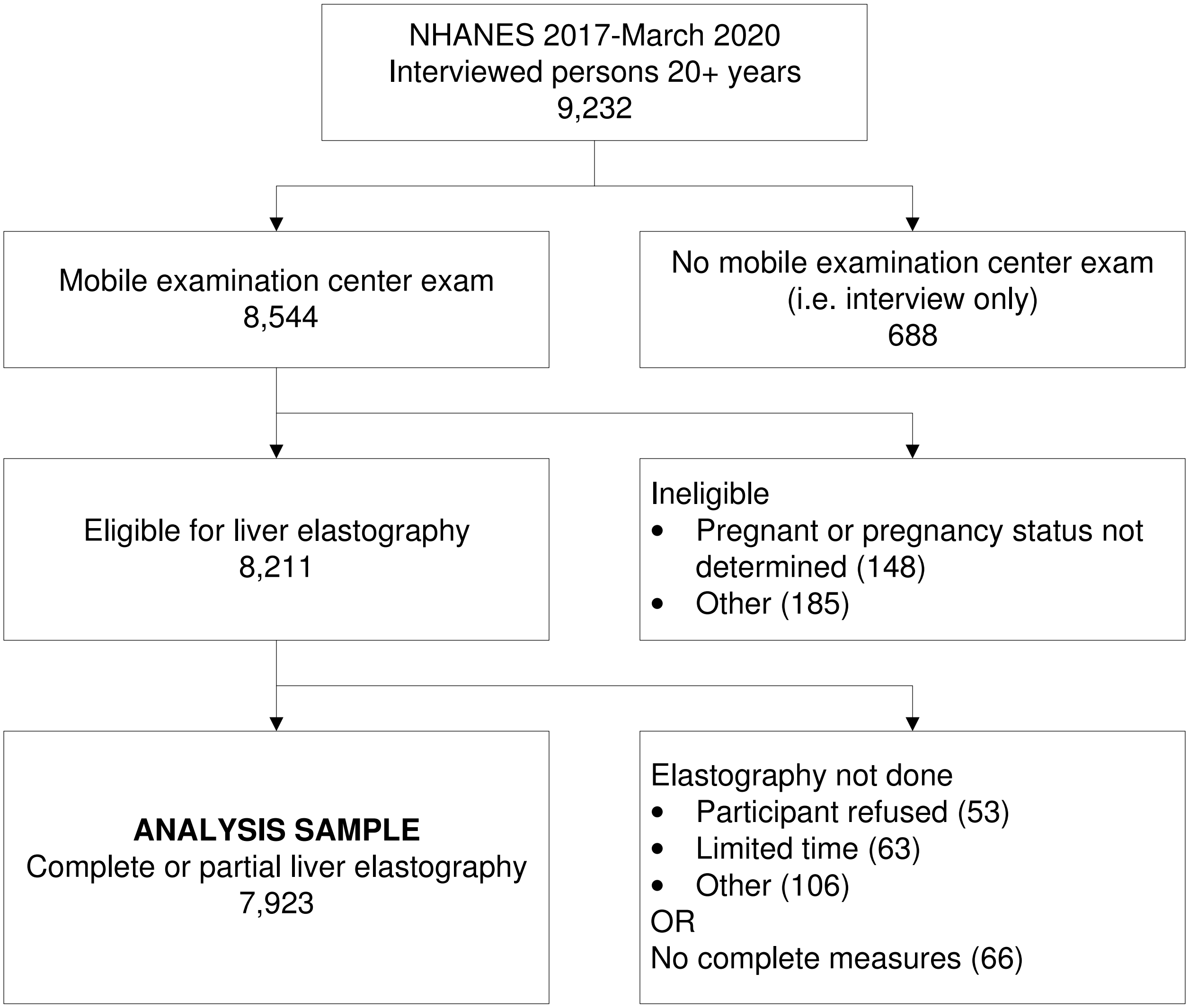

### Supplemental Figure 2.

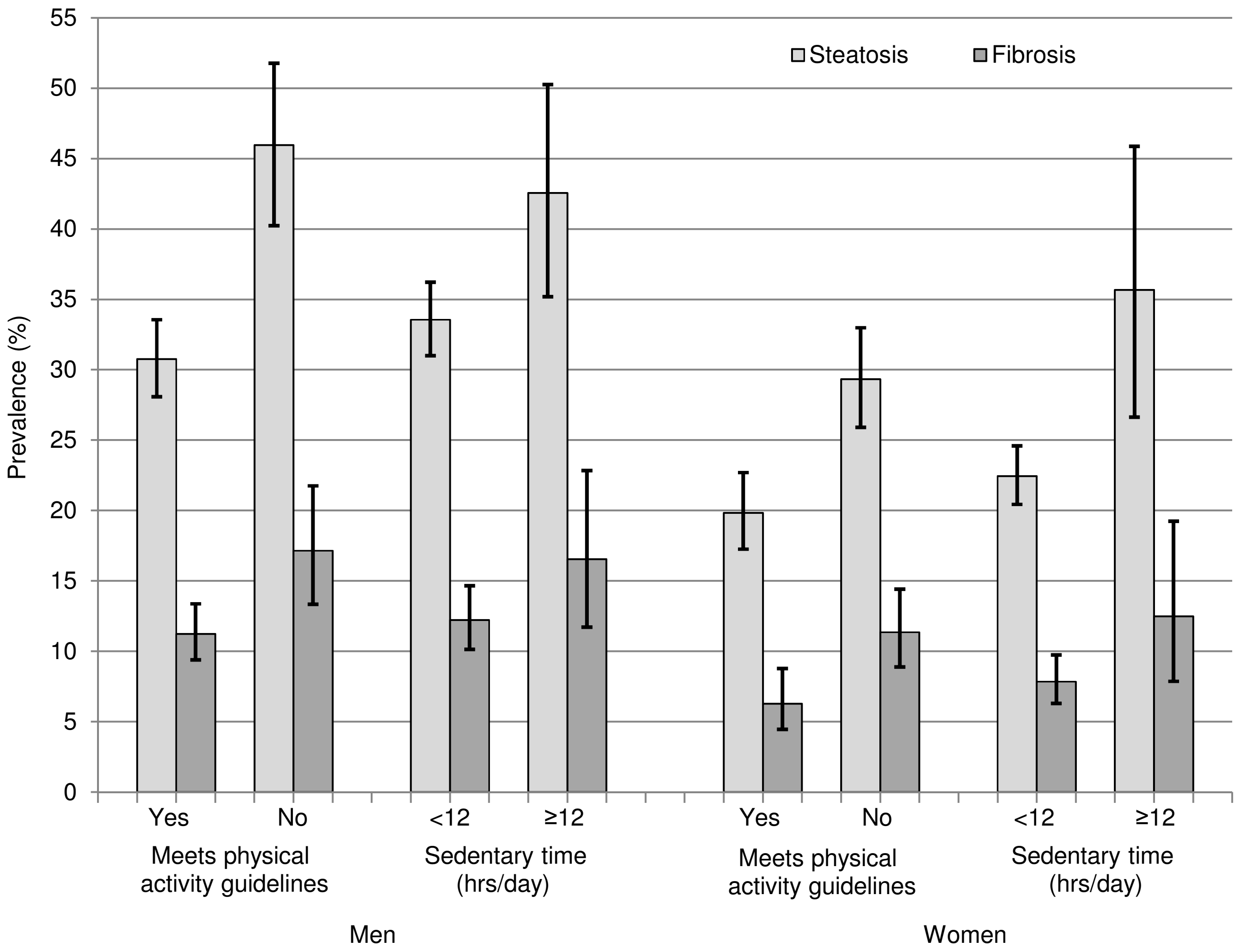

### Supplemental Figure 3.

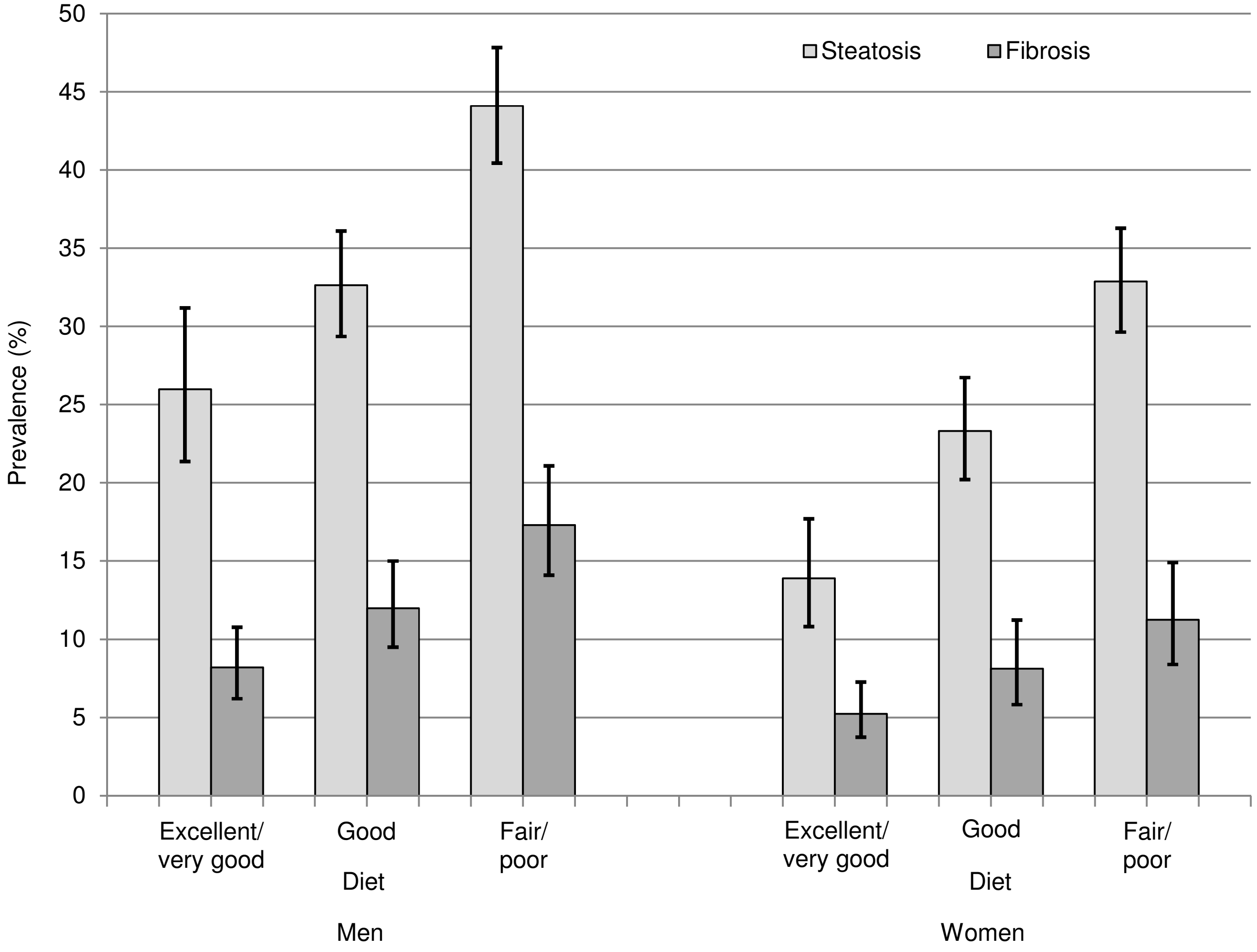

### Supplemental Figure 4.

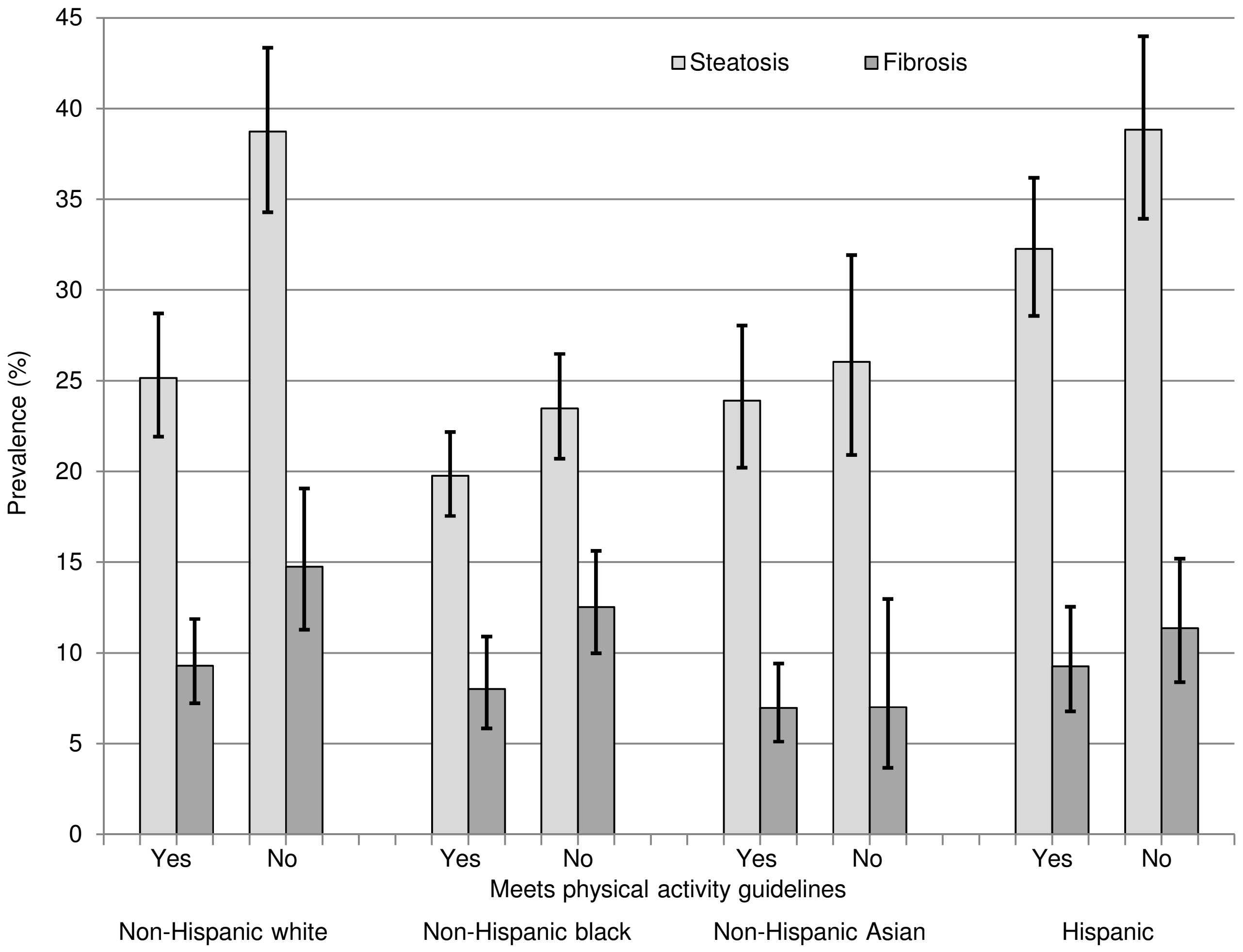

### Supplemental Figure 5.

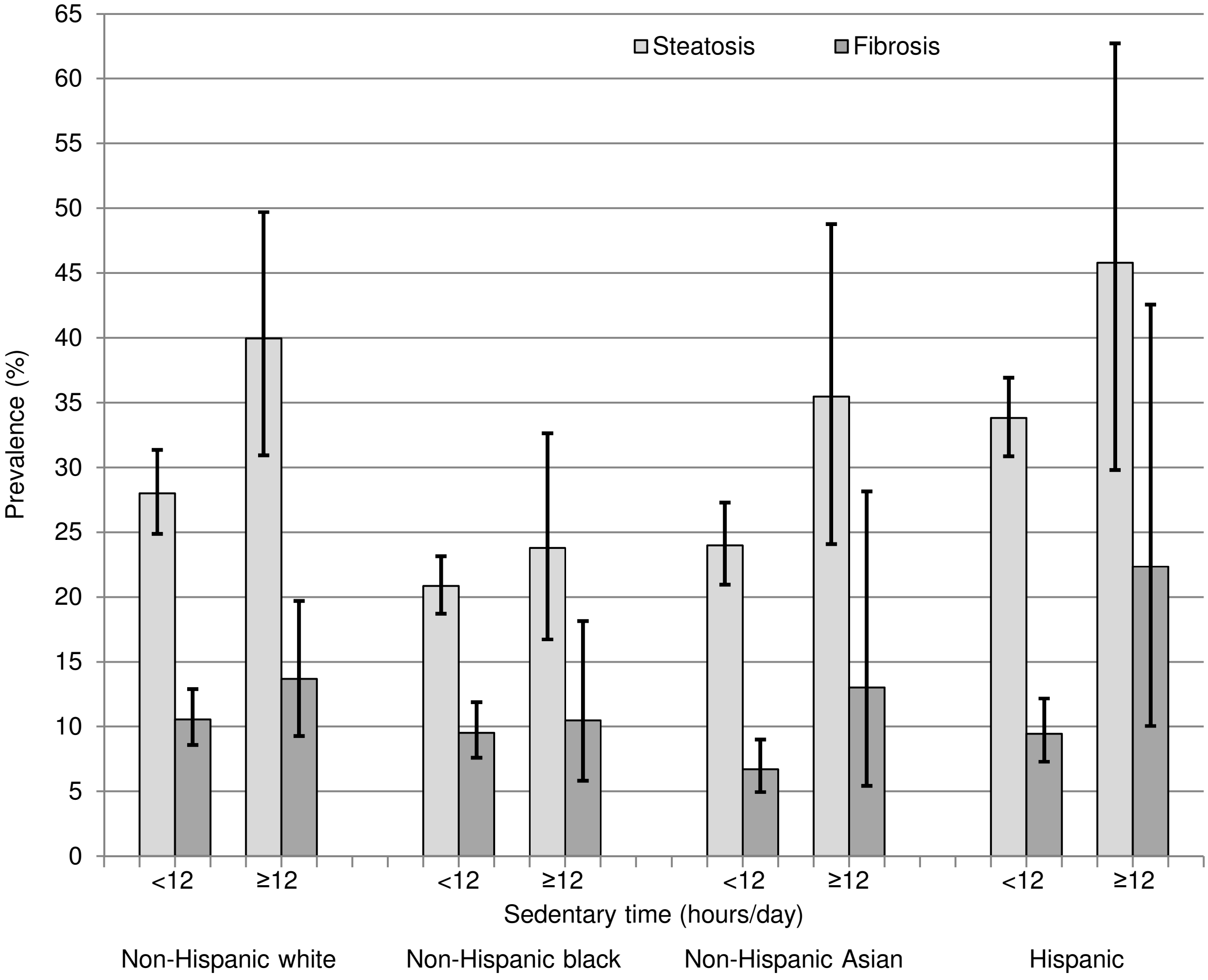

### Supplemental Figure 6.

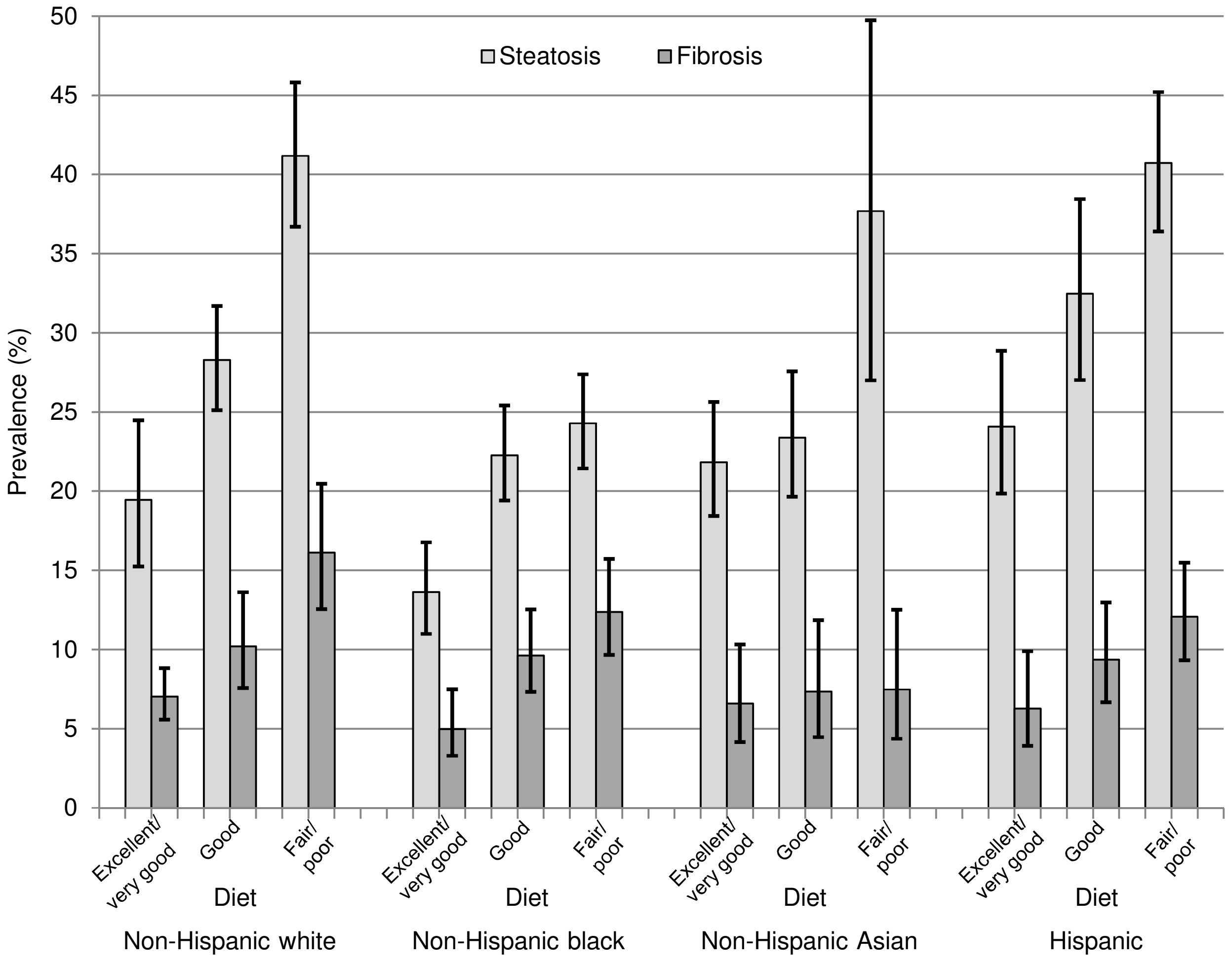

### Supplemental Figure 7.

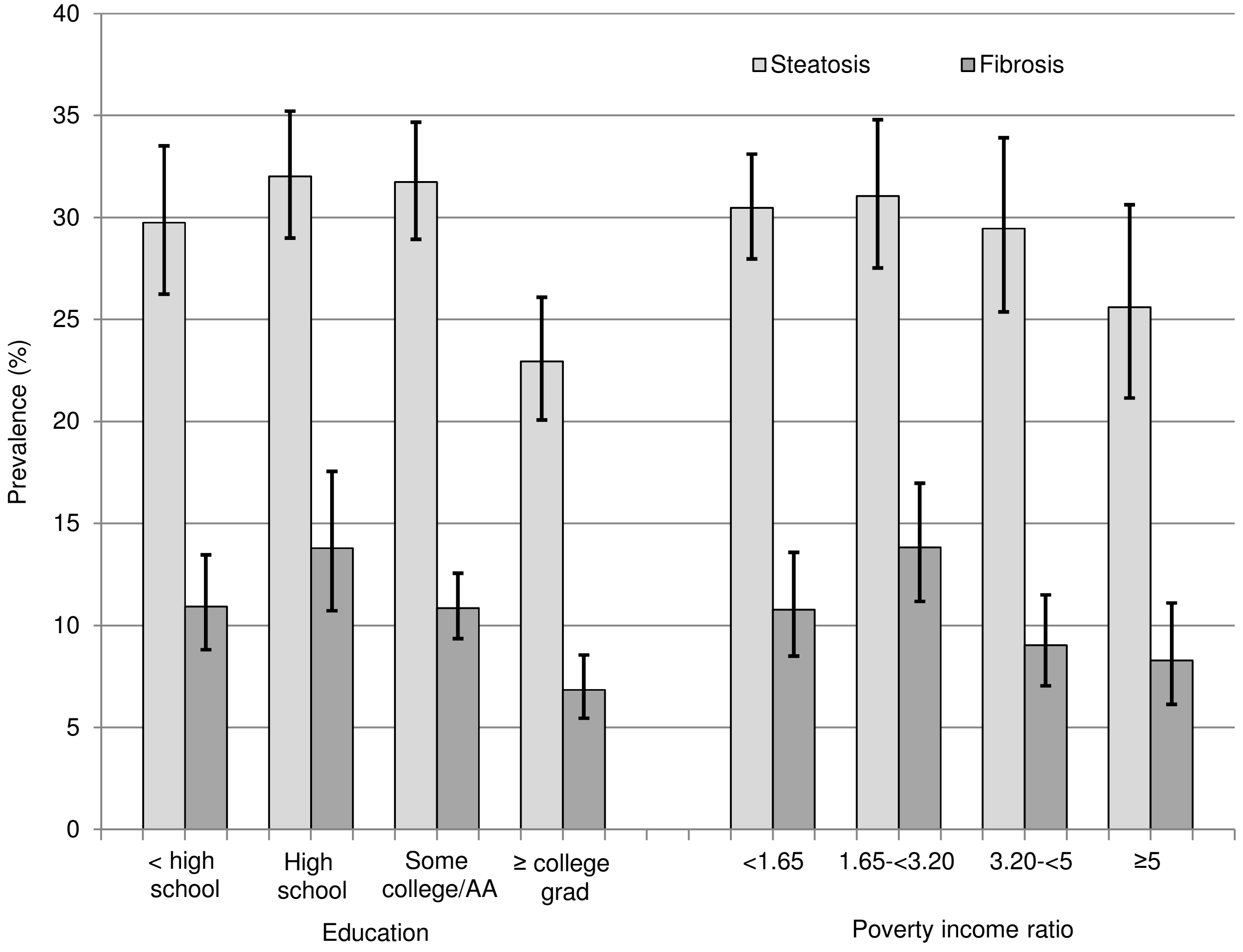

### Supplemental Figure 8.

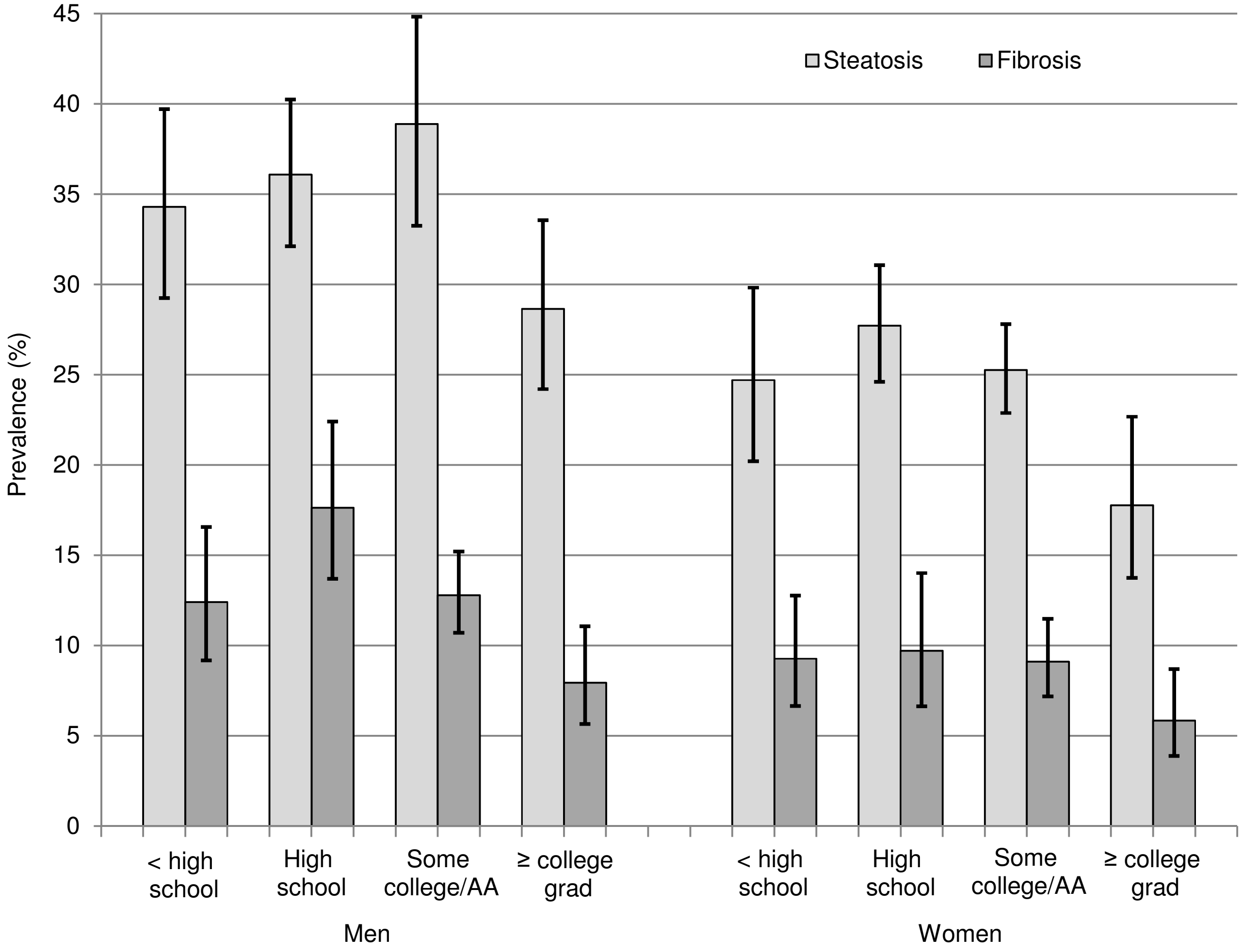

### Supplemental Figure 9.

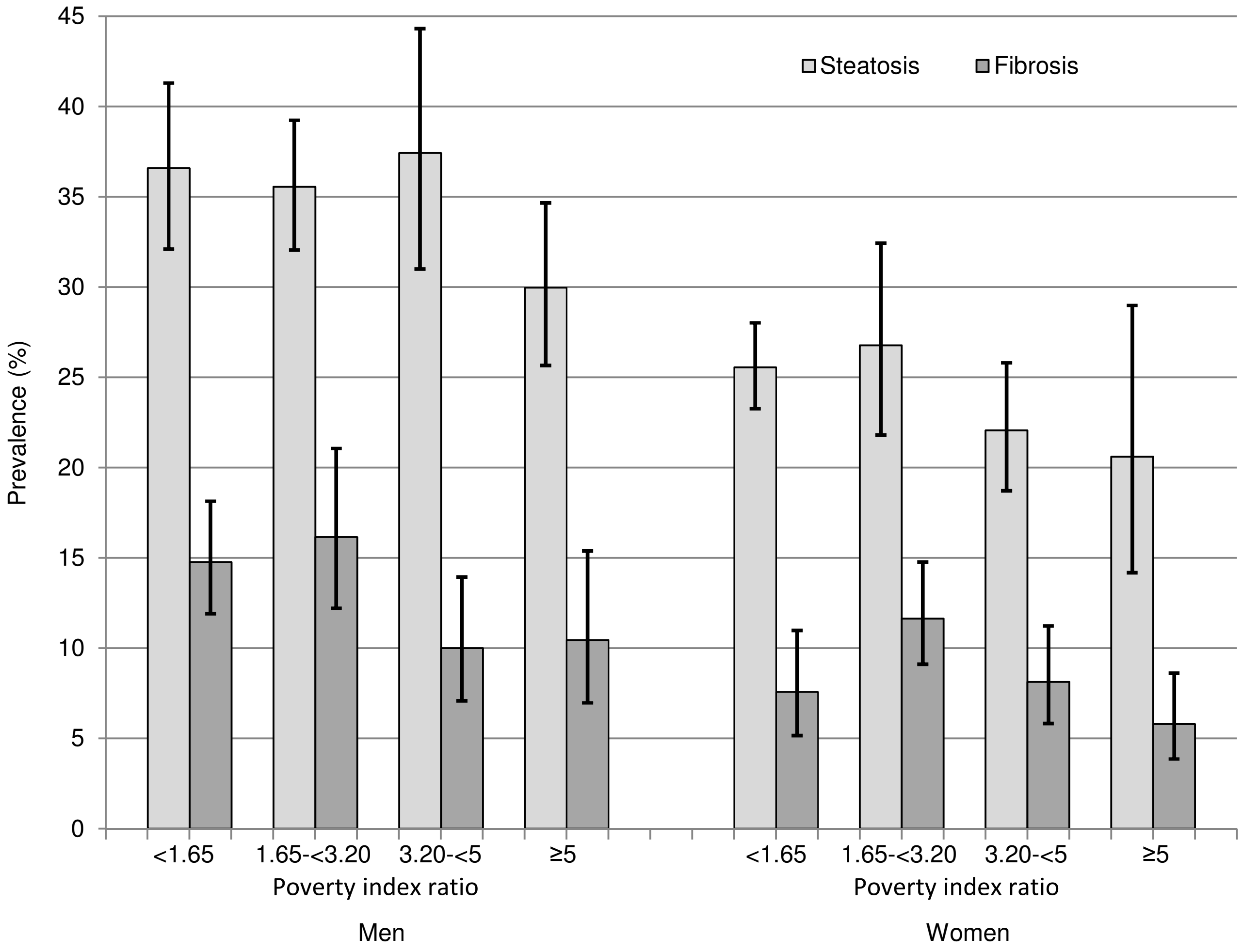

### Supplemental Figure 10.

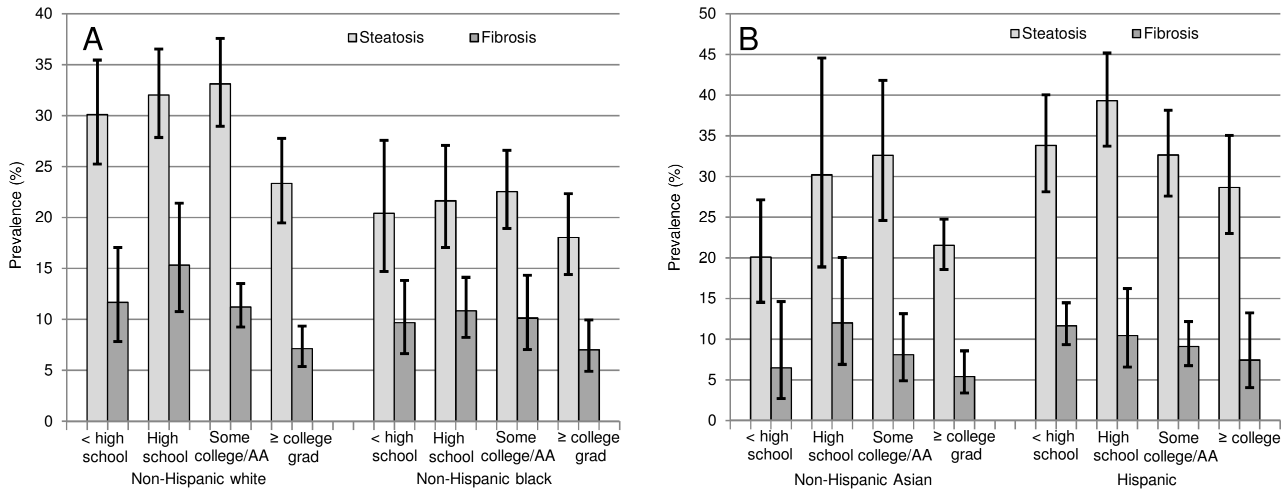

### Supplemental Figure 11.

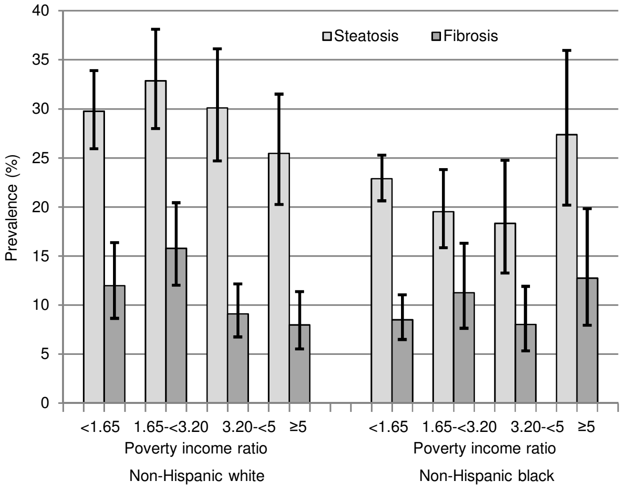

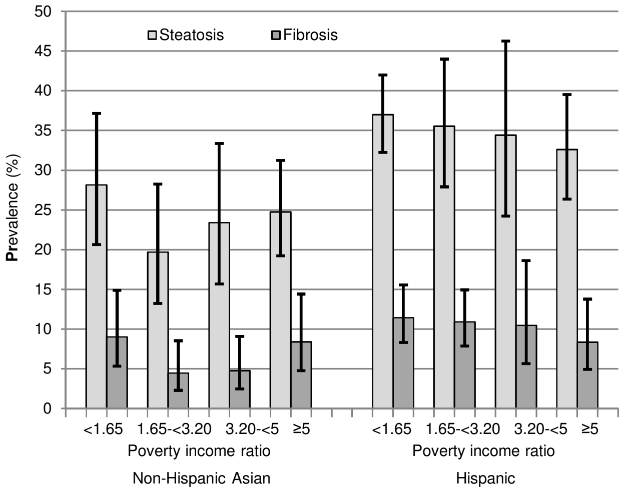
